# Pre-illness Clonal Hematopoiesis of Indeterminate Potential is Associated with Increased Morbidity and Mortality in Sepsis

**DOI:** 10.64898/2026.04.14.26350864

**Authors:** Nathaniel K. Berg, V. Eric Kerchberger, Yash Pershad, Robert W. Corty, Alexander G. Bick, Lorraine B. Ware

## Abstract

**Purpose:** Sepsis is a life-threatening syndrome causing significant morbidity and mortality, especially in older adults. Clonal hematopoiesis of indeterminate potential (CHIP) is an age-related condition linked to chronic illness and all-cause mortality. Here, we evaluated the association of pre-illness CHIP with mortality and morbidity in patients admitted to the ICU with sepsis.

**Methods:** We performed a retrospective study using a de-identified electronic health record linked with a DNA biorepository. We identified adult patients with sepsis who had DNA collected prior to ICU admission. We tested the association between CHIP status determined from whole-genome sequencing, and ICU mortality, organ support-free days, and long-term survival adjusting for age, sex, race and Sequential Organ Failure Assessment (SOFA) score on ICU admission.

**Results:** The cohort comprised 220 patients with pre-illness CHIP and 3,010 patients without. Pre-illness CHIP was associated with increased sepsis mortality (OR = 1.54, 95% CI 1.13 to 2.07, P = 0.005) and fewer days alive and free of organ support (-1.7 days, 95% CI-3.2 to-0.2, P = 0.028) after adjusting for age, sex, race, and SOFA score. In sepsis survivors, CHIP was also associated with increased long-term mortality after discharge (HR 1.40, 95% CI 1.01 to 1.93, P = 0.041).

**Conclusions:** Pre-illness CHIP was independently associated with increased mortality and morbidity in critically-ill adults with sepsis. These findings suggest that CHIP is a risk factor for sepsis severity. Uncovering the mechanism behind this link could lead to new treatments for sepsis, especially for the aging population.

## Introduction

Sepsis is a dysregulated host response to infection resulting in organ dysfunction and is associated with substantial morbidity and mortality [1]. Decades of research have advanced our understanding of the mechanisms underlying sepsis-induced organ injury and dysfunction, but improvements in mortality have been modest and are largely attributable to earlier recognition and increased adherence to best-practice interventions[2]. Furthermore, overall sepsis mortality is increasing, a trend likely driven by the rising number of at-risk individuals including the elderly and those with multiple comorbidities[3, 4].

Advanced age is recognized as risk factor for severity of sepsis, in part due to this population’s frailty and burden of comorbid conditions[5]. However, emerging evidence suggests that elderly patients also exhibit distinct biological responses to sepsis, including attenuated endothelial cell activation and dysregulated inflammatory signaling[6–8]. Identifying the factors that underlie these altered physiological responses in older adults is therefore critical to the development of targeted therapies to reduce the burden of sepsis-related morbidity and mortality in this high-risk population.

Clonal hematopoiesis of indeterminant potential (CHIP) occurs when somatic mutations in hematopoietic stem cells confer a clonal enrichment, resulting in detectable mutant blood cell populations at a variant allele fraction of 2% or greater, in the absence of overt hematologic malignancy or clonal disorder[9]. CHIP is associated with increased risk for numerous diseases including cardiovascular disease, autoimmune conditions, chronic kidney disease, malignancy and all-cause mortality[10, 11]. The prevalence of CHIP increases exponentially with age with prevalence of around 5% in patients 60 years of age, but 30% in individuals 80 years or older[12, 13]. A growing body of evidence from animals and humans suggests that CHIP may contribute to disease risk via dysregulated expression of inflammatory cytokines, impaired neutrophil function, and enhanced monocyte-endothelial interactions[14–18].

In this study, we examined the relationship between pre-illness CHIP and sepsis outcomes using a de-identified electronic health record database linked to a DNA bio-repository. Given the association between CHIP and aberrant inflammatory responses, we hypothesized that patients with CHIP would experience heightened risk of adverse outcomes in sepsis. Additionally, we tested whether presence of CHIP correlated with altered blood-cell counts during sepsis, if its presence in sepsis survivors is associated with higher mortality, and whether sepsis accelerates the growth of CHIP mutations in sepsis survivors.

## Methods

### Study Population

We studied adult patients enrolled in BioVU, Vanderbilt’s DNA biobank linked to a de-identified longitudinal electronic health record (EHR). All samples were generated from excess blood collected during routine medical care[19]. We identified patients with sepsis using either of two methods: 1) by applying a validated EHR algorithm[20] and 2) identifying the presence of an explicit ICD-9-CM and ICD-10-CM code in patients admitted to the ICU[21, 22] (codes available in online supplement **Table S1**). Library preparation and whole genome sequencing for approximately 250,000 BioVU participants was previously performed as part of an institutional program[23]. Presence of CHIP was classified as having a mutation in a CHIP driver gene with variant allele fraction (VAF) of 2% or more, consistent with established methods[24].

We selected patients who had DNA collected between two days and two years prior to hospital admission. This timeframe was implemented to ensure temporal proximity between DNA isolation and the ICU admission. A two-day lead-in period was used to ensure temporal precedence between CHIP status and ICU outcomes. Patients who had a concurrent or pre-existing diagnosis of myeloid neoplasm based on ICD-9-CM or ICD-10-CM codes were excluded (**Table S2**). We also excluded patients who were transferred out of ICU within 24 hours of admission unless they died in the first 24 hours. As a non-septic control group, we identified patients without sepsis who also met the above criteria and were admitted to a non-medical ICU (trauma, surgical, cardiovascular, and neurological ICUs).

## Outcomes

The primary outcome was in-hospital mortality, defined as death during the sepsis hospitalization or death occurring within two weeks of discharge to hospice to account for patients who were transitioned to an inpatient palliative care unit or to a “hospice-in-place” program which was catalogued as a discharge and re-admission to a separate hospice encounter in our EHR. We also tested the association between CHIP and in-hospital mortality among patients admitted to an ICU without sepsis to assess if the association was specific to sepsis, and if the effects of CHIP on mortality varied based on CHIP burden as measured by VAF or CHIP driver mutation.

Secondary outcomes included measures of organ support including 28-day vasopressor-free days, ventilator-free days, dialysis-free days, and total organ support-free days, as calculated using previously published methods[25]. Days requiring vasopressors, mechanical ventilation and dialysis were determined using medication records, ICD-10-PCS codes, and CPT procedure codes during each hospital stay (**Supplemental Tables E3 and E4**). Dialysis days were specifically calculated as the difference between the first and last date of dialysis events to reflect that some patients received intermittent dialysis and not continuous renal replacement therapy, and all dialysis analyses excluded patients with chronic renal failure requiring maintenance dialysis (ICD-9-CM and ICD-10-CM codes used listed in **Table S5**). Additional exploratory outcomes included leukocyte counts and differentials from the first 24-hours after ICU admission, long-term survival after discharge in sepsis survivors as ascertained from data available in the BioVU research EHR (**Supplementary Methods**), and longitudinal changes in CHIP mutation VAFs in a subset of BioVU participants who had longitudinal CHIP sequencing available from at least one time point before a sepsis ICU admission and at least one time point after discharge[26].

ICD-9-CM and ICD-10-CM codes during each hospital admission were used to generate Elixhauser comorbidity scores[27]. Vital signs and clinical laboratory data including maximum creatinine, minimum platelets, and maximum bilirubin from the first 24 hours of ICU admission were used to calculate sequential organ failure assessment (SOFA) scores[28].

## Statistical Analysis

Statistical analyses and figure generation were performed using R (version 4.5.1). Continuous variables were compared using the Wilcoxon rank-sum test and reported as medians with interquartile ranges, while categorical variables were assessed using Chi-squared or Fisher’s exact test. Differences in Elixhauser comorbidities were evaluated using logistic regression with an adjustment for age. Associations between pre-illness CHIP and outcomes including mortality, organ support-free days, and leukocyte counts were analyzed using logistic or linear regressions, adjusting for sex, race, initial SOFA score, and age at time of admission. We also tested for interaction effects between CHIP status and initial SOFA score and age at time of admission on mortality.

Additional Methods can be found in the online supplemental materials.

## Results

### Patient Characteristics

A total of 10,731 ICU patients with sepsis had available whole-genome sequencing. Among these, there were 3,230 patients meeting inclusion and exclusion criteria. Among them, 220 (6.8%) patients had CHIP mutations (**Figure 1**). In the non-sepsis cohort, 7,123 patients had available sequencing data, 2,121 met inclusion and exclusion criteria, and 130 (6.1%) had CHIP mutations (**Figure 1**). As expected, CHIP prevalence increased exponentially with age, with 5% CHIP prevalence at age 60 years, and 30% prevalence at age 90 years (**Figure S1**).

**Figure 1.**
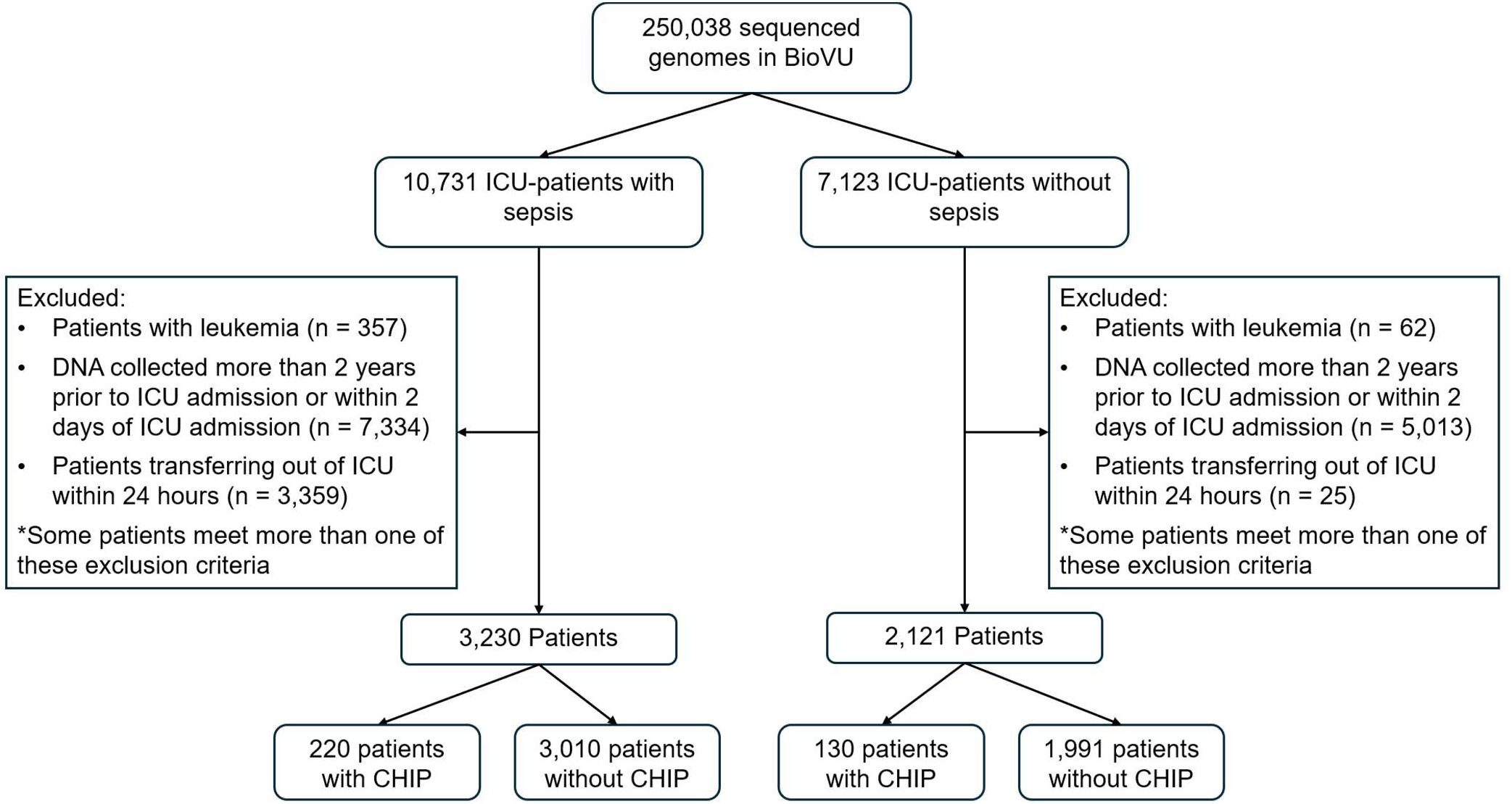
Study flow-diagram.

In the sepsis cohort, there were no significant differences between sex or admitting ICU between patients with and without CHIP (**Table 1**). As expected, patients with CHIP were older than those without. There was a trend towards increased prevalence of CHIP in white compared to black patients which was likely explained, in part, by older age among white patients in our cohort (median: 61 vs. 53 years; p<0.001). CHIP patients also had slightly lower body mass index (BMI). There were no significant differences in heart rate; respiratory parameters; or creatinine, bilirubin, or platelet levels at ICU admission based on CHIP status. While patients with CHIP did have a lower systolic blood pressure than those without, there was no difference in need for vasopressor support and there were no differences in initial SOFA scores. After adjusting for age, there were also no major differences in Elixhauser comorbidities at the time of hospital admission (**Table S6**). In the non-sepsis cohort, patients with CHIP were also older than those without, but there were overall no significant differences between patient characteristics and comorbidities (**Tables S7 and S8**).

**Table 1.**
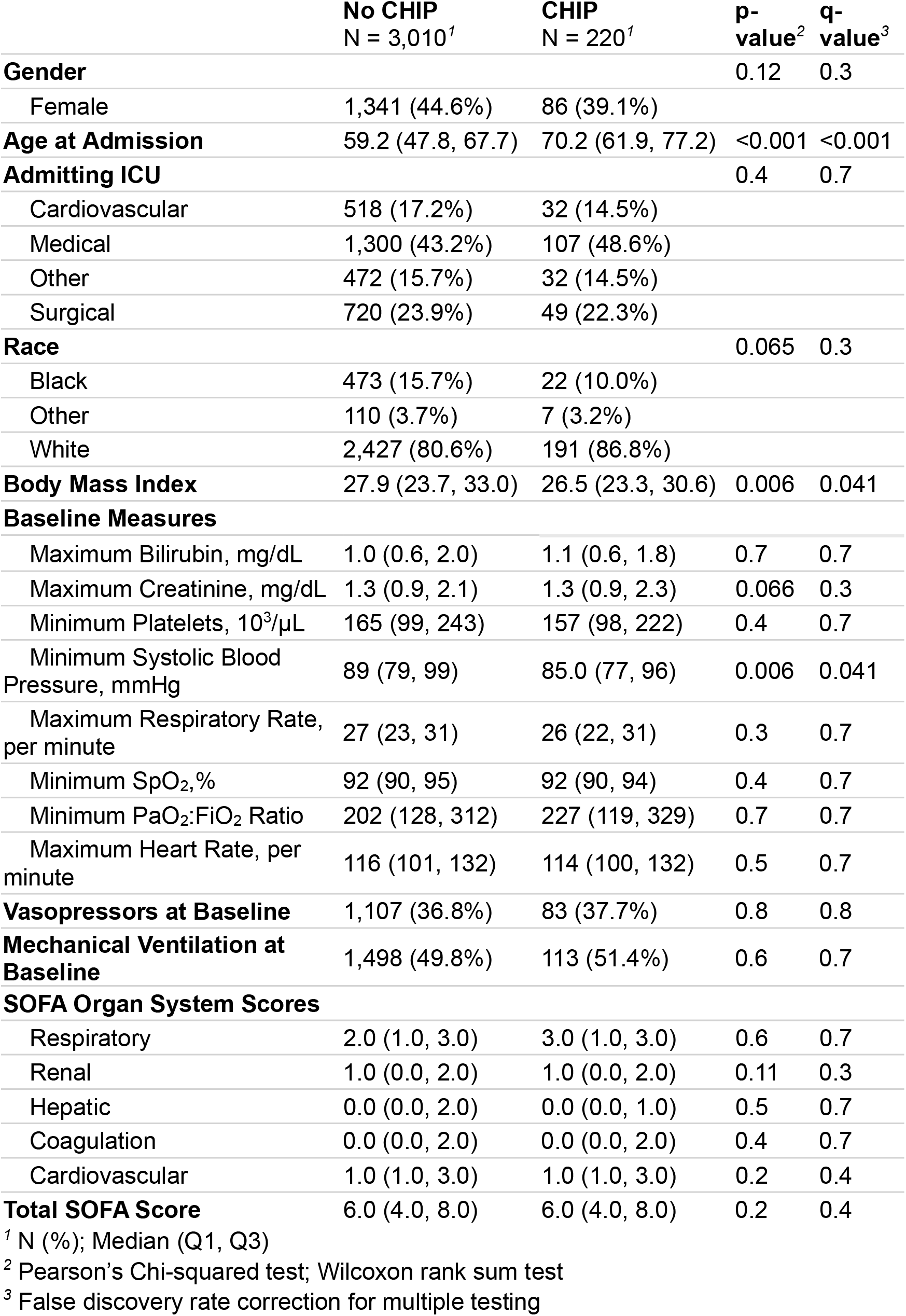
ICU sepsis cohort patient characteristics.

### Association of pre-illness CHIP with in-hospital mortality

Pre-illness CHIP was associated with increased in-hospital mortality (OR 1.54, 95% CI: 1.13 – 2.07, P = 0.005, **Figure 2**) after adjusting for age, sex, race, and initial SOFA score. There was no interaction between CHIP status and age (P> 0.9), suggesting the association of CHIP with mortality was consistent across all ages (**Table S9**). Furthermore, there was also no interaction between CHIP status and initial SOFA scores on sepsis mortality (P > 0.9, **Table S9**).

**Figure 2.**
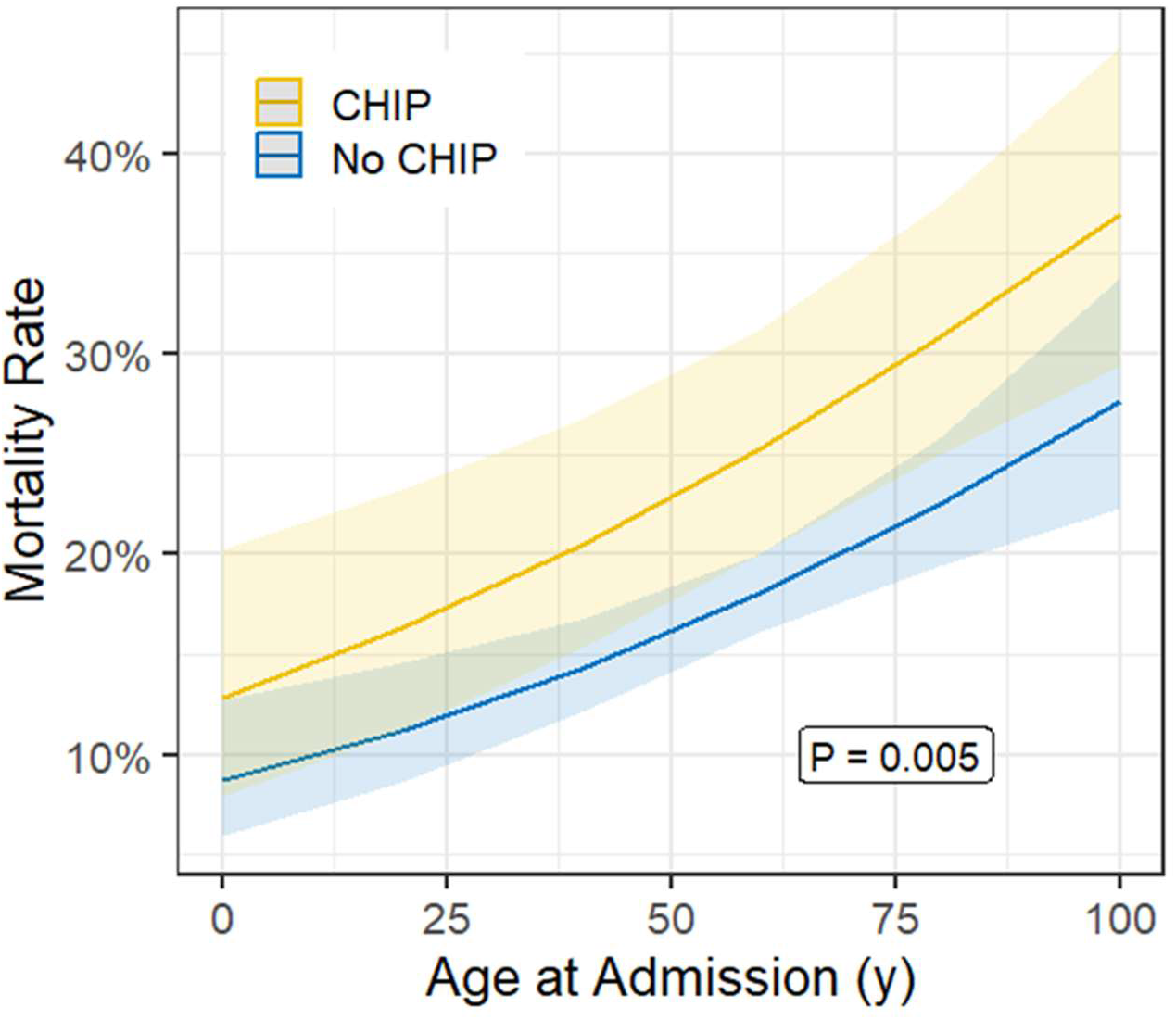
Inpatient mortality in sepsis patients admitted to the intensive care unit. Predicted mortality by logistic regression for patients with and without clonal hematopoiesis of indeterminant potential (CHIP) after adjusting for sex, race, age at time of admission, and baseline sequential organ failure assessment (SOFA) scores. Shading represents 95% confidence intervals.

Sensitivity analyses testing the effect of our DNA sampling window relative to ICU admission revealed similar associations between CHIP and in-hospital mortality when our DNA collection window was restricted to 10 days to two years (OR 1.44, 95% CI: 1.02 – 2.0, P = 0.035) or one year to two years (OR 2.1, 95% CI: 1.1 – 3.93, P = 0.022) prior to hospital admission, suggesting that sampling bias did not impact the association of CHIP with in-hospital mortality **(Figure S2)**. This association appeared specific to sepsis as CHIP status was not associated with in-hospital mortality among the patients who were admitted to a non-medical ICU and did not have sepsis (OR 0.62, 95% CI: 0.18–1.59, P = 0.4, **Figure S3)**. Additionally, while previous studies have shown that CHIP burden as measured by VAF or the dominant driver mutation can exert a dose-dependent effect on long-term disease development, we did not observe any significant differences in hospital mortality among patients with CHIP based on either VAF or driver mutation (**Figure S4**) [12, 13, 29].

### Pre-illness CHIP is associated with fewer organ support-free days

After adjusting for age, sex, race, and initial SOFA score, patients with pre-illness CHIP had fewer organ support-free days compared to those without CHIP (1.7 fewer days, 95% CI: 3.2 – 0.3 fewer days, P = 0.018, **Figure 3**). This included fewer days free from assisted ventilation (1.8 fewer days, 95% CI: 3.2 – 0.28 fewer days, P = 0.02), vasopressors (1.7 fewer days, 95% CI: 3.1 – 0.3 fewer days, P = 0.017), and renal replacement therapy (3 fewer days, 95% CI: 5.3 – 0.74 fewer days, P = 0.009, **Figure 3**).

**Figure 3.**
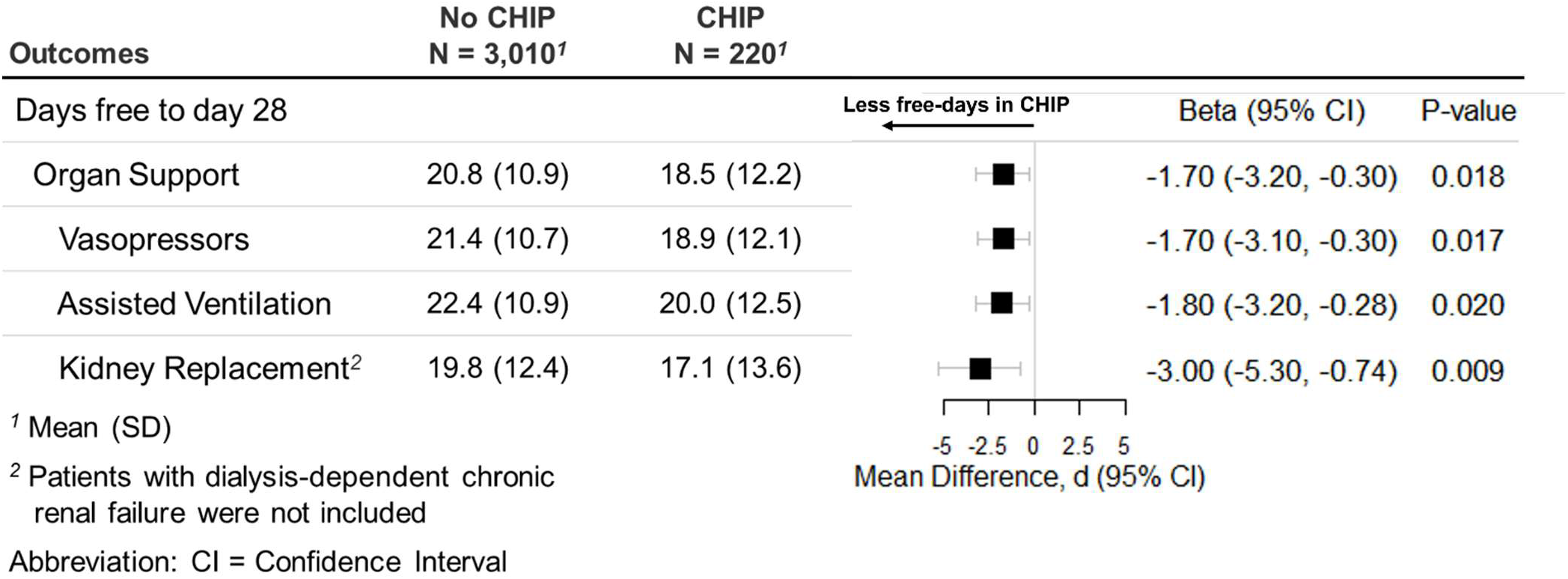
Clonal hematopoiesis of indeterminant potential (CHIP) is associated with fewer organ support-free days. Organ support-free days were calculated as days free of vasopressors, assisted ventilation, or dialysis. Dialysis-free days were not calculated in patients already undergoing dialysis for chronic kidney disease. The data are presented as mean days and standard deviation (SD). Forest plot displays the mean difference in days (CHIP versus no CHIP). Beta values are adjusted for age, sex, race and SOFA score based on linear regression with 95% confidence intervals (CI). P values are based on Wald’s t-test.

### Pre-illness CHIP is associated with altered peripheral hematologic profiles during the sepsis event

Total leukocyte counts were higher at all time points in patients with CHIP versus no CHIP (mean increase of 2.8 x 10^3^ cells/uL; 95% CI: 1.5 – 4.0 x 10^3^ cells/uL; P < 0.001, **Figure S5A**). This was largely driven by higher neutrophil counts (mean increase of 2.8 x 10^3^ cells/uL in patients with CHIP; 95% CI: 1.7 – 3.9 x 10^3^ cells/uL; P < 0.001), with a trend towards increased neutrophil percentages (mean increase of 2% in patients with CHIP’ 95% CI: 0% – 4%’ P = 0.082, **Figure S5B, C**). While there was no difference in absolute monocyte counts (**Figure S5D**), patients with CHIP had lower monocyte percentages (mean decrease of 1%; 95% CI-2% – 0%; P = 0.019; **Figure S5E**). Patients with CHIP also had lower absolute lymphocyte counts (mean decrease of 0.16 x 10^3^ cells/uL; 95% CI:-0.31 –-0.01 x 10^3^ cells/uL; P = 0.038), without a difference in lymphocyte percentages (**Figure S5F, G**). There were no differences in eosinophil counts or percentages (**Figure S5H, I**). These hematologic profile differences appeared to be specifically related to the acute sepsis event as we observed no difference in pre-sepsis cell count profiles in the 2,105 patients (149 [7.1%] with CHIP) who had outpatient leukocyte count and differentials performed within 60 days of DNA collection for CHIP sequencing (**Figure S6**).

### Sepsis survivors with CHIP have worse long-term survival

Among 2,577 patients (156 [6.1%] with CHIP) who survived their initial critical illness event, CHIP was associated with worse one-year (55% vs. 75%, P < 0.001), two-year (46% vs. 69%, P = 0.049), and five-year survival (39% vs. 60%, P = 0.037, **Table S10**). This association remained significant after adjusting for age at admission, sex, race and initial SOFA score on ICU admission (HR 1.40, 95% CI: 1.01 – 1.93, P = 0.041; **Figure 4**).

**Figure 4.**
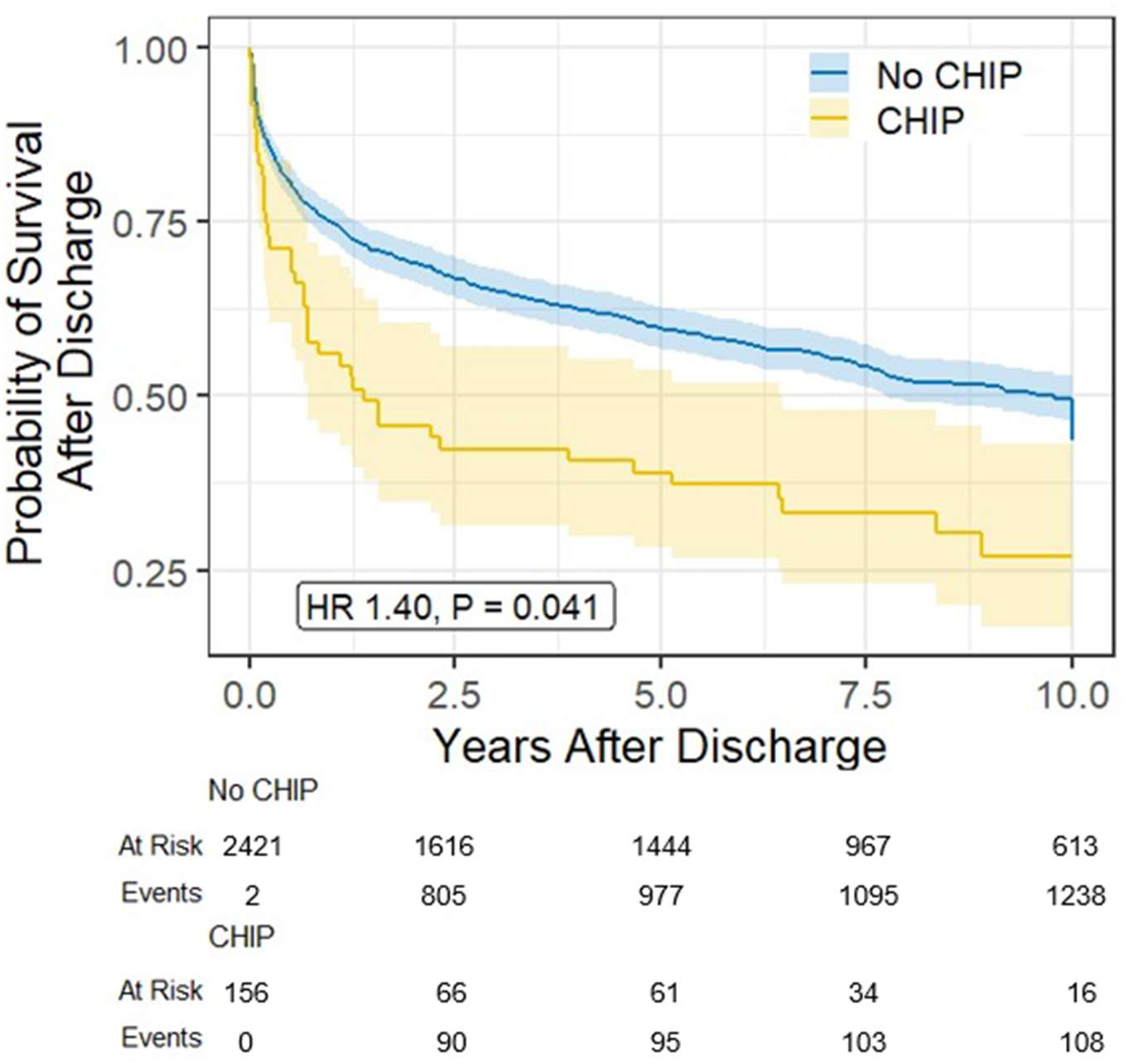
Clonal hematopoiesis of indeterminant potential (CHIP) in sepsis survivors is associated with increased long-term mortality. The survival curve represents 156 patients with CHIP and 2421 without. The hazard ratio (HR) was adjusted for age at time of admission, sex, race, and initial sequential organ failure assessment (SOFA) score using Cox regression analysis.

### Sepsis accelerates the growth of CHIP allele fractions

As an exploratory analysis to test if sepsis affected the kinetics of somatic CHIP mutation accumulation, we identified 11 BioVU patients who had CHIP sequencing performed between 2 years and 2 days before their ICU sepsis admission and subsequently had a second DNA sample sequenced between 2 weeks and 1 year after their admission and compared them to BioVU participants with longitudinal CHIP sequencing who were never hospitalized between CHIP measurements (**Figure S7A**). After matching for sex, age at first sample, race, initial VAF, gene mutation, mutation type, and time interval between the two samples (**Table S11**), we observed an accelerated rate of growth in CHIP VAF among sepsis survivors (+9% per year, 95% CI: 1% - 17%, P = 0.024, **Figure S7B**) compared to never-sepsis controls. To test whether post-sepsis shifts in myeloid and granulocyte counts drove the observed CHIP VAF growth, we analyzed cell counts and differentials from both DNA collection time points. Patients with interval sepsis admissions showed an increase in neutrophil counts at the second time point compared to controls, though this difference did not reach statistical significance (10.0 x 10^3^ cells/uL vs. 4.0 x 10^3^ cells/uL, P=0.093, **Table S12**). Similarly, we observed a non-significant trend toward decreased relative lymphocyte counts in CHIP patients both before and after sepsis admissions (**Table S12**). Thus, while persistent neutrophil expansion may explain VAF growth, a larger sample size is needed to confirm these results.

## Discussion

To our knowledge, this study is the first to establish a significant association between CHIP and outcomes in critically-ill adults with sepsis. CHIP was independently associated with prolonged organ support and increased in-hospital mortality across all age groups and baseline SOFA scores. Furthermore, the impact of CHIP extended beyond the period of acute illness, as sepsis survivors with CHIP had worse long-term survival and an acceleration in CHIP clonal expansion. Collectively, these data underscore the importance of CHIP as a marker of a particularly vulnerable patient population in both the ICU and post-ICU care settings.

While prior literature has linked CHIP to elevated sepsis risk, existing studies are limited by highly selected cohorts and a reliance on longitudinal incidence in healthy populations, leaving the acute clinical impact of CHIP in real-world sepsis patients unaddressed. In a recent secondary analysis of the Aspirin in Reducing Events in the Elderly (ASPREE) trial, community-dwelling adults who had CHIP clones with higher mutational burden (VAF ≥10%) had increased risk for sepsis and sepsis related deaths, while those with low-burden clones (VAF <10%) did not.[30] Similarly, an analysis of 453,510 individuals with CHIP genotyping in the UK Biobank reported a 27% increase in sepsis incidence among individuals with CHIP, much of which was driven by mediation through incident leukemias[30]. Both the ASPREE trial and the UK Biobank study focused on community-dwelling individuals and assessed the risk of *incident* sepsis during longitudinal follow up. Conversely, we evaluate patients with *prevalent* sepsis and leveraged the high-density clinical data available in BioVU’s research EHR to interrogate how CHIP modulates the acute response to infection and affects long-term outcomes in survivors. Additionally, the ASPREE trial restricted enrollment to individuals aged 70 years or older but were free of cardiovascular disease, anemia, dementia, or life-limiting illnesses; whereas the UK Biobank study was restricted to adults aged 40-70 years. Consequently, the ASPREE cohort was significantly older, yet healthier and less comorbid than our hospital-based biobank population, while the UK Biobank had a narrower age range. ASPREE utilized CHIP-specific sequencing technology, while both the UK Biobank and our study used existing whole-genome sequencing data for CHIP ascertainment which may have affected our ability to detect low-abundance CHIP clones and limited our ability to test associations with VAF burden. The UK Biobank study also relied upon ICD codes for identifying sepsis which has a lower sensitivity and increased risk for misclassification than prospective phenotyping by clinical investigators used in the ASPREE trial or EHR-based phenotyping used in our study.[31] Lastly, while the UK Biobank study reported that most of the observed association between CHIP and sepsis was mediated through incident leukemias, our study specifically excluded patients with prevalent or incident hematologic malignancy suggesting our results were not driven by cancer development.

Although previous studies established a VAF ≥10% as the threshold for increased risk of several diseases [12, 32], VAF was not associated with in-hospital mortality in this cohort. As CHIP VAF was measured before sepsis onset, the lack of VAF dependence in our study may stem from unobserved clonal changes occurring between DNA sampling and ICU admission. Alternatively, acute clonal expansion driven by systemic inflammation and stress hematopoiesis during sepsis may be clinically more relevant than pre-hospital VAF[33–36]. Confirming this hypothesis would require serial DNA collection and CHIP VAF measurements throughout the course of sepsis in a future study.

Our results also demonstrated an altered circulating white blood cell profile in sepsis patients with CHIP characterized by elevated absolute neutrophil counts and reduced absolute lymphocyte counts. These findings are consistent with existing literature establishing that neutrophilia and lymphopenia during sepsis are associated with increased organ dysfunction and death[37, 38]. Neutrophils act as primary effectors in acute inflammation, where an exacerbated response drives tissue destruction, subsequent organ injury, and coagulopathy[39]. Furthermore, increased granulopoiesis during sepsis is associated with immunosuppression, more severe disease, and early mortality[40]. While our findings do not provide direct evidence for dysregulated function of white blood cells in patients with CHIP, our data support a hypothesis that CHIP mutations may influence sepsis mortality through effects on the function of descendant leukocytes.

Lastly, our findings suggest that critical illness due to sepsis may accelerate the expansion of CHIP clones, potentially heightening the risk of known CHIP-associated diagnoses and overall all-cause mortality[10, 12, 29, 32]. This aligns with prior findings that hospital admissions for infections and HIV increase the risk of developing CHIP, supporting the theory that infection and inflammation fuel CHIP pathogenesis[41, 42]. Additionally, people with IL6R mutations show slower long-term growth of TET2 clones, suggesting that IL-6 activity helps drive CHIP expansion[43]. Although our findings support the link between sepsis-related inflammation and subsequent CHIP expansion, our small sample size precludes a definitive conclusion. Larger-sized longitudinal studies will be essential to validate this finding.

This study has several strengths. We used a large and well-established institutional EHR biobank with over 20 years of longitudinal data, providing the statistical power necessary to study a relatively uncommon genetic exposure. An additional strength is that we were able to precisely correlate pre-illness CHIP status with sepsis outcomes. Unlike most critical-care genetic studies that rely on DNA collected at the time of or during ICU admission, the longitudinal data and biospecimens in BioVU allowed assessment of pre-illness risk. Another strength is the use of validated EHR algorithms that have higher sensitivity than claims data alone to identify patients with sepsis, reducing the risk of misclassification[20].

There are also some limitations. First, our study is limited by the lack of an independent validation cohort with deep phenotyping comparable to BioVU. UK Biobank lacks inpatient vital signs, laboratory data, and critical care tracking, making it impossible to adjust for acute illness severity as we performed in our study. While the *All of Us* Research Program Biobank does include inpatient clinical data, at the time of this writing, it exhibits a relatively smaller EHR follow up time due to its more recent inception and lower rate of CHIP genotyping availability.[24] Second, data collection from a single quaternary referral center inherently enriches the patient population for sicker individuals, which may limit generalizability of our results to broader populations. Third, since we relied on institutional EHR data, information on hospitalizations at other centers not linked to our EHR is unavailable. Fourth, while the DNA biospecimens used were not collected during the acute sepsis encounter, they could have been obtained during other periods of illness. While DNA in BioVU is generally collected during outpatient encounters, it is not fully discernable if patients were at baseline health at the collection time. Additionally, some patients could have developed CHIP in the time between pre-illness DNA collection and ICU admission, resulting in misclassification bias. Finally, assessment of CHIP in this cohort was based on approximately 35X sequencing depth rather than the “gold standard” 1000X coverage of deep sequencing (longitudinal CHIP sequencing data), which could lead to an underestimation of CHIP and effect sizes[44].

In conclusion, pre-illness CHIP in critically ill adults with sepsis was associated with higher in-hospital mortality, fewer organ support-free days, and worse long-term survival. Furthermore, CHIP was associated with altered hematologic profiles during sepsis, suggesting a maladaptive inflammatory response, and sepsis was associated with increased expansion of mutant CHIP clones in longitudinal follow up. These findings indicate that CHIP may serve as a critical biological mechanism driving poorer sepsis outcomes, particularly within the vulnerable aged population. As genomic data become more integrated into clinical care, detecting CHIP could improve risk stratification and long-term management for critically ill patients.

## Data Availability

All data produced in the present study are available upon reasonable request to the authors

## Acknowledgments

We extend our sincere gratitude to the BioVU participants for their invaluable voluntary contributions to this research. The whole-genome sequencing (WGS) of 250,000 individuals was made possible through the Alliance for Genomic Discovery (AGD), a collaborative initiative funded by NashBio, Illumina, and a consortium of industry partners including Amgen, AbbVie, AstraZeneca, Bayer, BMS, GSK, Merck, and Novo Nordisk. Sequencing was executed at deCODE genetics utilizing Illumina’s sequencing technology. The BioVU projects at Vanderbilt University Medical Center are supported by numerous sources: institutional funding, private agencies, and federal grants. These include NIH funded Shared Instrumentation Grant S10OD017985, S10RR025141, and S10OD025092; and CTSA grants UL1TR002243, UL1TR000445, and UL1RR024975 from the National Center for Advancing Translational Sciences.

## Availability of data and materials

The datasets use and/or analyzed in this study are available from the corresponding author on reasonable request

## Declarations

### Ethics approval and consent to participate

The data reproduced in this article utilized DNA sequencing that was procured via BioVU (our institutional BioBank), which provides de-identified samples. This study was reviewed and deemed exempt by our Institutional Review Board. The BioBank protocols are in accordance with the ethical standards of our institution and with the 1964 Helsinki declaration and its later amendments or comparable ethical standards

### Consent for publication

Not applicable

### Author Contributions

Study conception and design: N.K.B, V.E.K, A.G.B, L.B.W. Data collection: V.E.K, R.W.C., Y.P.

Data analysis: N.K.B.

Writing of the manuscript: N.K.B.

Revising critically for important intellectual content and final approval: all authors.

### Take-Home Message

This study establishes pre-existing clonal hematopoiesis of indeterminate potential (CHIP) as an independent risk factor for increased mortality and morbidity in septic ICU patients, regardless of age or illness severity. These findings suggest CHIP drives worse sepsis outcomes in aging populations and could reveal novel therapeutic insights.

This article has an online data supplement.

