## Supplemental Data for "Pre-illness Clonal Hematopoiesis of Indeterminate Potential is Associated with Increased Morbidity and Mortality in Sepsis"

Online Data Supplement

Table of Contents


| Content | Page |
|---|---|
| **Supplemental Methods** | 2 |
| **Supplemental Tables** | |
| Table S1. ICD 9/10 CM codes used for identification of patients with sepsis admitted to the ICU. | 4 |
| Table S2. ICD 9/10 CM codes used for identification of patients with leukemia. | 6 |
| Table S3. CPT, ICD-10-PCS, and ICD-9-Proc codes used to calculate assisted ventilation days. | 13 |
| Table S4. CPT, ICD-10-PCS, and ICD-9-Proc codes used to calculate dialysis days. | 14 |
| Table S5. ICD 9/10 codes used to identify patients with end stage renal disease. | 15 |
| Table S6. ICU sepsis patient cohort Elixhauser comorbidities. | 16 |
| Table S7. ICU non-sepsis cohort patient characteristics | 17 |
| Table S8. ICU non-sepsis patient cohort Elixhauser comorbidities. | 18 |
| Table S9. Logistic regression model results for odds of inpatient mortality in critically-ill sepsis patients. | 19 |
| Table S10. Survival probability in sepsis survivors with and CHIP. | 20 |
| Table S11. Case-Control matching results for variant allele fraction growth analysis. | 21 |
| Table S12. Blood counts and differentials matched to first and second specimen collections used to measure CHIP variant allele fraction growth. | 22 |
| **Supplemental Figures** | |
| Figure S1. Distribution of CHIP fraction across age in patients admitted to the ICU. | 23 |
| Figure S2. Sensitivity analysis of ICU sepsis mortality by DNA collection timing. | 24 |
| Figure S3. Inpatient mortality in non-sepsis patients admitted to non-medical intensive care units. | 25 |
| Figure S4. Association of CHIP variant allele frequency and specific CHIP gene mutation with hospital mortality in sepsis patients admitted to the ICU. | 26 |
| Figure S5. Association of CHIP with cell-count differentials. | 27 |
| Figure S6. Leukocyte counts and differentials proximal to the time of pre-illness biospecimen collection. | 28 |
| Figure S7. CHIP variant allele fraction expansion accelerates in sepsis patients who survive an ICU admission. | 29 |
| **Supplemental References** | 30 |

### Supplemental Methods

#### Assessment of hematologic profiles

As CHIP results in the preferential expansion of myeloid lineages[1], we characterized its influence on the peripheral leukocyte profiles of patients with sepsis. We extracted peripheral leukocyte counts and differential data collected during routine clinical care from hospital admission through the first five days in the ICU. For baseline leukocyte assessments, we analyzed the laboratory values closest to the DNA collection date, ensuring it was recorded prior to admission and no more than 60 days away from the DNA collection.

#### Long-term survival

We evaluated long-term post-sepsis survival using EHR mortality data. BioVU aggregates death records from four primary sources: (i) in-hospital mortality at our medical center; (ii) out-of-hospital deaths reported to our Medical Records Operations by family members, external facilities, clinicians, or administrative staff; (iii) institutional disease registries; (iv) the Social Security Death Index (SSDI), available through December 31, 2016; and deaths identified from obituary and claims data provided through Datavant (New York, New York, USA). For individuals missing a specific death date (restricted to a subset of SSDI records), the date of last contact served as a proxy, defined by the final recorded diagnosis code, clinical visit, or electronic communication. Patients without a documented death record were assumed alive and censored at their date of last contact.

Long-term survival probabilities at one-, two-, and five-year timepoints were compared using a Z-score test and an overall hazard ratio for mortality was determined by Cox regression using the *Survival* R package, with age at time of admission, race, and sex included as covariates.

**Longitudinal CHIP clonal dynamics**

Prior studies suggest that IL-6 signaling drives the growth of CHIP variant alleles[2]. Consequently, we hypothesized that the inflammation driven by sepsis may act as a catalyst for CHIP VAF expansion. To investigate this, we leveraged previously published longitudinal CHIP data from a subset of 3,000 individuals in BioVU with DNA samples collected at two points in time and sequenced with a deep, targeted sequencing assay[3, 4]. As above, we selected individuals whose first blood draw occurred between two years and two days prior to an ICU admission for sepsis; patients also had to have a follow-up sample that was collected between two weeks and one-year post-admission. These intervals were chosen to more specifically capture CHIP expansion driven by sepsis-related inflammation rather than age-related expansion. Control subjects were patients in BioVU who had not been admitted to the ICU with sepsis and did not have ICD codes for leukemia. We performed case-control matching using the R package *MatchIt*. Control patients were matched (in a 2:1 ratio) first by exact CHIP gene and mutation type (e.g. missense, frameshift, nonsense, etc.) followed by nearest-neighbor matching for sex, age at first DNA collection, initial VAF, and the time interval between samples. CHIP growth rates were calculated based on a exponential growth formula as previously described[3].

Table S1. ICD 9/10 CM codes used for identification of patients with sepsis admitted to the ICU.

| Code | Version | Description |
|---|---|---|
| A02.1 | ICD10CM | Salmonella sepsis |
| A20.0 | ICD10CM | Bubonic plague |
| A20.7 | ICD10CM | Septicemic plague |
| A22.7 | ICD10CM | Anthrax sepsis |
| A26.7 | ICD10CM | Erysipelothrix sepsis |
| A32.7 | ICD10CM | Listerial sepsis |
| A40.0 | ICD10CM | Sepsis due to streptococcus, group A |
| A40.1 | ICD10CM | Sepsis due to streptococcus, group B |
| A40.3 | ICD10CM | Sepsis due to Streptococcus pneumoniae |
| A40.8 | ICD10CM | Other streptococcal sepsis |
| A40.9 | ICD10CM | Streptococcal sepsis, unspecified |
| A41.01 | ICD10CM | Sepsis due to Methicillin susceptible Staphylococcus aureus |
| A41.02 | ICD10CM | Sepsis due to Methicillin resistant Staphylococcus aureus |
| A41.1 | ICD10CM | Sepsis due to other specified staphylococcus |
| A41.2 | ICD10CM | Sepsis due to unspecified staphylococcus |
| A41.3 | ICD10CM | Sepsis due to Hemophilus influenzae |
| A41.4 | ICD10CM | Sepsis due to anaerobes |
| A41.50 | ICD10CM | Gram-negative sepsis, unspecified |
| A41.51 | ICD10CM | Sepsis due to Escherichia coli [E. coli] |
| A41.52 | ICD10CM | Sepsis due to Pseudomonas |
| A41.53 | ICD10CM | Sepsis due to Serratia |
| A41.54 | ICD10CM | Sepsis due to Acinetobacter baumannii |
| A41.59 | ICD10CM | Other Gram-negative sepsis |
| A41.81 | ICD10CM | Sepsis due to Enterococcus |
| A41.89 | ICD10CM | Other specified sepsis |
| A41.9 | ICD10CM | Sepsis, unspecified organism |
| A42.7 | ICD10CM | Actinomycotic sepsis |
| A54.86 | ICD10CM | Gonococcal sepsis |
| B37.7 | ICD10CM | Candidal sepsis |
| O03.37 | ICD10CM | Sepsis following incomplete spontaneous abortion |
| O03.87 | ICD10CM | Sepsis following complete or unspecified spontaneous abortion |
| O04.87 | ICD10CM | Sepsis following (induced) termination of pregnancy |
| O07.37 | ICD10CM | Sepsis following failed attempted termination of pregnancy |
| O08.82 | ICD10CM | Sepsis following ectopic and molar pregnancy |
| O85 | ICD10CM | Puerperal sepsis |
| O86.04 | ICD10CM | Sepsis following an obstetrical procedure |
| P36.0 | ICD10CM | Sepsis of newborn due to streptococcus, group B |
| P36.10 | ICD10CM | Sepsis of newborn due to unspecified streptococci |
| P36.19 | ICD10CM | Sepsis of newborn due to other streptococci |
| P36.2 | ICD10CM | Sepsis of newborn due to Staphylococcus aureus |
| P36.30 | ICD10CM | Sepsis of newborn due to unspecified staphylococci |
| P36.39 | ICD10CM | Sepsis of newborn due to other staphylococci |
| P36.4 | ICD10CM | Sepsis of newborn due to Escherichia coli |
| P36.5 | ICD10CM | Sepsis of newborn due to anaerobes |

Table S1 Continued. ICD 9/10 CM codes used for identification of patients with sepsis admitted to the ICU.

| Code | Version | Description |
|---|---|---|
| P36.8 | ICD10CM | Other bacterial sepsis of newborn |
| P36.9 | ICD10CM | Bacterial sepsis of newborn, unspecified |
| R65.20 | ICD10CM | Severe sepsis without septic shock |
| R65.21 | ICD10CM | Severe sepsis with septic shock |
| T81.44XA | ICD10CM | Sepsis following a procedure, initial encounter |
| T81.44XD | ICD10CM | Sepsis following a procedure, subsequent encounter |
| T81.44XS | ICD10CM | Sepsis following a procedure, sequela |
| 003.1 | ICD9CM | Salmonella septicemia |
| 020.2 | ICD9CM | Septicemic plague |
| 022.3 | ICD9CM | Anthrax septicemia |
| 038 | ICD9CM | Septicemia |
| 038.0 | ICD9CM | Streptococcal septicemia |
| 038.1 | ICD9CM | Staphylococcal septicemia |
| 038.10 | ICD9CM | Staphylococcal septicemia, unspecified |
| 038.11 | ICD9CM | Methicillin susceptible Staphylococcus aureus septicemia |
| 038.12 | ICD9CM | Methicillin resistant Staphylococcus aureus septicemia |
| 038.19 | ICD9CM | Other staphylococcal septicemia |
| 038.2 | ICD9CM | Pneumococcal septicemia [Streptococcus pneumoniae septicemia] |
| 038.3 | ICD9CM | Septicemia due to anaerobes |
| 038.4 | ICD9CM | Septicemia due to other gram-negative organisms |
| 038.40 | ICD9CM | Septicemia due to gram-negative organism, unspecified |
| 038.41 | ICD9CM | Septicemia due to hemophilus influenzae [H. influenzae] |
| 038.42 | ICD9CM | Septicemia due to escherichia coli [E. coli] |
| 038.43 | ICD9CM | Septicemia due to pseudomonas |
| 038.44 | ICD9CM | Septicemia due to serratia |
| 038.49 | ICD9CM | Other septicemia due to gram-negative organisms |
| 038.8 | ICD9CM | Other specified septicemias |
| 038.9 | ICD9CM | Unspecified septicemia |
| 054.5 | ICD9CM | Herpetic septicemia |
| 670.20 | ICD9CM | Puerperal sepsis, unspecified as to episode of care or not applicable |
| 670.22 | ICD9CM | Puerperal sepsis, delivered, with mention of postpartum complication |
| 670.24 | ICD9CM | Puerperal sepsis, postpartum condition or complication |
| 771.81 | ICD9CM | Septicemia [sepsis] of newborn |
| 785.52 | ICD9CM | Septic shock |
| 995.90 | ICD9CM | Systemic inflammatory response syndrome, unspecified |
| 995.91 | ICD9CM | Sepsis |
| 995.92 | ICD9CM | Severe sepsis |

Table S2. ICD 9/10 CM codes used for identification of patients with leukemia.

| Code | Version | Description |
|---|---|---|
| 202.4 | ICD9CM | Leukemic reticuloendotheliosis |
| 202.4 | ICD9CM | Leukemic reticuloendotheliosis, unspecified site, extranodal and solid organ sites |
| 202.41 | ICD9CM | Leukemic reticuloendotheliosis involving lymph nodes of head, face, and neck |
| 202.42 | ICD9CM | Leukemic reticuloendotheliosis involving intrathoracic lymph nodes |
| 202.43 | ICD9CM | Leukemic reticuloendotheliosis involving intra-abdominal lymph nodes |
| 202.44 | ICD9CM | Leukemic reticuloendotheliosis involving lymph nodes of axilla and upper limb |
| 202.45 | ICD9CM | Leukemic reticuloendotheliosis involving lymph nodes of inguinal region and lower limb |
| 202.46 | ICD9CM | Leukemic reticuloendotheliosis involving intrapelvic lymph nodes |
| 202.47 | ICD9CM | Leukemic reticuloendotheliosis involving spleen |
| 202.48 | ICD9CM | Leukemic reticuloendotheliosis involving lymph nodes of multiple sites |
| 203 | ICD9CM | Multiple myeloma and immunoproliferative neoplasms |
| 203 | ICD9CM | Multiple myeloma |
| 203 | ICD9CM | Multiple myeloma without mention of having achieved remission |
| 203.01 | ICD9CM | Multiple myeloma in remission |
| 203.02 | ICD9CM | Multiple myeloma in relapse |
| 203.1 | ICD9CM | Plasma cell leukemia |
| 203.1 | ICD9CM | Plasma cell leukemia without mention of having achieved remission |
| 203.11 | ICD9CM | Plasma cell leukemia in remission |
| 203.12 | ICD9CM | Plasma cell leukemia in relapse |
| 203.8 | ICD9CM | Other immunoproliferative neoplasms |
| 203.8 | ICD9CM | Other immunoproliferative neoplasms without mention of having achieved remission |
| 203.81 | ICD9CM | Other immunoproliferative neoplasms in remission |
| 203.82 | ICD9CM | Other immunoproliferative neoplasms in relapse |
| 204 | ICD9CM | Lymphoid leukemia |
| 204 | ICD9CM | Lymphoid leukemia, acute |
| 204 | ICD9CM | Acute lymphoid leukemia without mention of having achieved remission |
| 204.01 | ICD9CM | Acute lymphoid leukemia in remission |
| 204.02 | ICD9CM | Lymphoid leukemia, acute in relapse |
| 204.1 | ICD9CM | Lymphoid leukemia, chronic |
| 204.1 | ICD9CM | Chronic lymphoid leukemia without mention of having achieved remission |
| 204.11 | ICD9CM | Chronic lymphoid leukemia in remission |
| 204.12 | ICD9CM | Lymphoid leukemia, chronic in relapse |
| 204.2 | ICD9CM | Lymphoid leukemia, subacute |
| 204.2 | ICD9CM | Subacute lymphoid leukemia without mention of having achieved remission |
| 204.21 | ICD9CM | Subacute lymphoid leukemia in remission |
| 204.22 | ICD9CM | Lymphoid leukemia, subacute in relapse |
| 204.8 | ICD9CM | Other lymphoid leukemia |
| 204.8 | ICD9CM | Other lymphoid leukemia without mention of having achieved remission |
| 204.81 | ICD9CM | Other lymphoid leukemia in remission |
| 204.82 | ICD9CM | Other lymphoid leukemia in relapse |
| 204.9 | ICD9CM | Unspecified lymphoid leukemia |
| 204.9 | ICD9CM | Unspecified lymphoid leukemia without mention of having achieved remission |
| 204.91 | ICD9CM | Unspecified lymphoid leukemia in remission |

Table S2 Continued. ICD 9/10 CM codes used for identification of patients with leukemia.

| Code | Version | Description |
|---|---|---|
| 204.92 | ICD9CM | Unspecified lymphoid leukemia in relapse |
| 205 | ICD9CM | Myeloid leukemia |
| 205 | ICD9CM | Myeloid leukemia, acute |
| 205 | ICD9CM | Acute myeloid leukemia without mention of having achieved remission |
| 205.01 | ICD9CM | Acute myeloid leukemia in remission |
| 205.02 | ICD9CM | Myeloid leukemia, acute in relapse |
| 205.1 | ICD9CM | Myeloid leukemia, chronic |
| 205.1 | ICD9CM | Chronic myeloid leukemia without mention of having achieved remission |
| 205.11 | ICD9CM | Chronic myeloid leukemia in remission |
| 205.12 | ICD9CM | Myeloid leukemia, chronic in relapse |
| 205.2 | ICD9CM | Myeloid leukemia, subacute |
| 205.2 | ICD9CM | Subacute myeloid leukemia without mention of having achieved remission |
| 205.21 | ICD9CM | Subacute myeloid leukemia in remission |
| 205.22 | ICD9CM | Myeloid leukemia, subacute in relapse |
| 205.3 | ICD9CM | Myeloid sarcoma |
| 205.3 | ICD9CM | Myeloid sarcoma without mention of having achieved remission |
| 205.31 | ICD9CM | Myeloid sarcoma in remission |
| 205.32 | ICD9CM | Myeloid sarcoma in relapse |
| 205.8 | ICD9CM | Other myeloid leukemia |
| 205.8 | ICD9CM | Other myeloid leukemia without mention of having achieved remission |
| 205.81 | ICD9CM | Other myeloid leukemia in remission |
| 205.82 | ICD9CM | Other myeloid leukemia in relapse |
| 205.9 | ICD9CM | Unspecified myeloid leukemia |
| 205.9 | ICD9CM | Unspecified myeloid leukemia without mention of having achieved remission |
| 205.91 | ICD9CM | Unspecified myeloid leukemia in remission |
| 205.92 | ICD9CM | Unspecified myeloid leukemia in relapse |
| 206 | ICD9CM | Monocytic leukemia |
| 206 | ICD9CM | Monocytic leukemia, acute |
| 206 | ICD9CM | Acute monocytic leukemia without mention of having achieved remission |
| 206.01 | ICD9CM | Acute monocytic leukemia in remission |
| 206.02 | ICD9CM | Monocytic leukemia, acute in relapse |
| 206.1 | ICD9CM | Monocytic leukemia, chronic |
| 206.1 | ICD9CM | Chronic monocytic leukemia without mention of having achieved remission |
| 206.11 | ICD9CM | Chronic monocytic leukemia in remission |
| 206.12 | ICD9CM | Monocytic leukemia, chronic in relapse |
| 206.2 | ICD9CM | Monocytic leukemia, subacute |
| 206.2 | ICD9CM | Subacute monocytic leukemia without mention of having achieved remission |
| 206.21 | ICD9CM | Subacute monocytic leukemia in remission |
| 206.22 | ICD9CM | Monocytic leukemia, subacute in relapse |
| 206.8 | ICD9CM | Other monocytic leukemia |
| 206.8 | ICD9CM | Other monocytic leukemia without mention of having achieved remission |
| 206.81 | ICD9CM | Other monocytic leukemia in remission |
| 206.82 | ICD9CM | Other monocytic leukemia in relapse |
| 206.9 | ICD9CM | Unspecified monocytic leukemia |

Table S2 Continued. ICD 9/10 CM codes used for identification of patients with leukemia.

| Code | Version | Description |
|---|---|---|
| 206.9 | ICD9CM | Unspecified monocytic leukemia without mention of having achieved remission |
| 206.91 | ICD9CM | Unspecified monocytic leukemia in remission |
| 206.92 | ICD9CM | Unspecified monocytic leukemia in relapse |
| 207 | ICD9CM | Other specified leukemia |
| 207 | ICD9CM | Acute erythremia and erythroleukemia |
| 207 | ICD9CM | Acute erythremia and erythroleukemia without mention of having achieved remission |
| 207.01 | ICD9CM | Acute erythremia and erythroleukemia in remission |
| 207.02 | ICD9CM | Acute erythremia and erythroleukemia in relapse |
| 207.1 | ICD9CM | Chronic erythremia |
| 207.1 | ICD9CM | Chronic erythremia without mention of having achieved remission |
| 207.11 | ICD9CM | Chronic erythremia in remission |
| 207.12 | ICD9CM | Chronic erythremia in relapse |
| 207.2 | ICD9CM | Megakaryocytic leukemia |
| 207.2 | ICD9CM | Megakaryocytic leukemia without mention of having achieved remission |
| 207.21 | ICD9CM | Megakaryocytic leukemia in remission |
| 207.22 | ICD9CM | Megakaryocytic leukemia in relapse |
| 207.8 | ICD9CM | Other specified leukemia |
| 207.8 | ICD9CM | Other specified leukemia without mention of having achieved remission |
| 207.81 | ICD9CM | Other specified leukemia in remission |
| 207.82 | ICD9CM | Other specified leukemia in relapse |
| 208 | ICD9CM | Leukemia of unspecified cell type |
| 208 | ICD9CM | Leukemia of unspecified cell type, acute |
| 208 | ICD9CM | Acute leukemia of unspecified cell type without mention of having achieved remission |
| 208.01 | ICD9CM | Acute leukemia of unspecified cell type in remission |
| 208.02 | ICD9CM | Leukemia of unspecified cell type, acute in relapse |
| 208.1 | ICD9CM | Leukemia of unspecified cell type, chronic |
| 208.1 | ICD9CM | Chronic leukemia of unspecified cell type without mention of having achieved remission |
| 208.11 | ICD9CM | Chronic leukemia of unspecified cell type in remission |
| 208.12 | ICD9CM | Leukemia of unspecified cell type, chronic in relapse |
| 208.2 | ICD9CM | Leukemia of unspecified cell type, subacute |
| 208.2 | ICD9CM | Subacute leukemia of unspecified cell type without mention of having achieved remission |
| 208.21 | ICD9CM | Subacute leukemia of unspecified cell type in remission |
| 208.22 | ICD9CM | Leukemia of unspecified cell type, subacute in relapse |
| 208.8 | ICD9CM | Other leukemia of unspecified cell type |
| 208.8 | ICD9CM | Other leukemia of unspecified cell type without mention of having achieved remission |
| 208.81 | ICD9CM | Other leukemia of unspecified cell type in remission |
| 208.82 | ICD9CM | Other leukemia of unspecified cell type in relapse |
| 208.9 | ICD9CM | Unspecified leukemia |
| 208.9 | ICD9CM | Unspecified leukemia without mention of having achieved remission |
| 208.91 | ICD9CM | Unspecified leukemia in remission |
| 208.92 | ICD9CM | Leukemia in relapse NOS |
| V10.6 | ICD9CM | Personal history of leukemia |
| V10.60 | ICD9CM | Personal history of unspecified leukemia |

Table S2 Continued. ICD 9/10 CM codes used for identification of patients with leukemia.

| Code | Version | Description |
|---|---|---|
| V10.61 | ICD9CM | Personal history of lymphoid leukemia |
| V10.62 | ICD9CM | Personal history of myeloid leukemia |
| V10.63 | ICD9CM | Personal history of monocytic leukemia |
| V10.69 | ICD9CM | Personal history of other leukemia |
| C91.01 | ICD10CM | Acute lymphoblastic leukemia, in remission |
| C91.02 | ICD10CM | Acute lymphoblastic leukemia, in relapse |
| C91.0 | ICD10CM | Acute lymphoblastic leukemia [ALL] |
| C91.00 | ICD10CM | Acute lymphoblastic leukemia not having achieved remission |
| C92.11 | ICD10CM | Chronic myeloid leukemia, BCR/ABL-positive, in remission |
| C92.10 | ICD10CM | Chronic myeloid leukemia, BCR/ABL-positive, not having achieved remission |
| C92.12 | ICD10CM | Chronic myeloid leukemia, BCR/ABL-positive, in relapse |
| C92.1 | ICD10CM | Chronic myeloid leukemia, BCR/ABL-positive |
| C92.Z0 | ICD10CM | Other myeloid leukemia not having achieved remission |
| C92.32 | ICD10CM | Myeloid sarcoma, in relapse |
| C92.2 | ICD10CM | Atypical chronic myeloid leukemia, BCR/ABL-negative |
| C92.22 | ICD10CM | Atypical chronic myeloid leukemia, BCR/ABL-negative, in relapse |
| C92.Z2 | ICD10CM | Other myeloid leukemia, in relapse |
| C92.90 | ICD10CM | Myeloid leukemia, unspecified, not having achieved remission |
| D46 | ICD10CM | Myelodysplastic syndromes |
| C92.3 | ICD10CM | Myeloid sarcoma |
| C92.20 | ICD10CM | Atypical chronic myeloid leukemia, BCR/ABL-negative, not having achieved remission |
| C92.Z | ICD10CM | Other myeloid leukemia |
| C92.Z1 | ICD10CM | Other myeloid leukemia, in remission |
| C92.92 | ICD10CM | Myeloid leukemia, unspecified in relapse |
| C92.9 | ICD10CM | Myeloid leukemia, unspecified |
| C92.31 | ICD10CM | Myeloid sarcoma, in remission |
| C92.21 | ICD10CM | Atypical chronic myeloid leukemia, BCR/ABL-negative, in remission |
| C92.30 | ICD10CM | Myeloid sarcoma, not having achieved remission |
| C92 | ICD10CM | Myeloid leukemia |
| C92.91 | ICD10CM | Myeloid leukemia, unspecified in remission |
| C95.0 | ICD10CM | Acute leukemia of unspecified cell type |
| Z85.6 | ICD10CM | Personal history of leukemia |
| C94.30 | ICD10CM | Mast cell leukemia not having achieved remission |
| C95 | ICD10CM | Leukemia of unspecified cell type |
| C94.20 | ICD10CM | Acute megakaryoblastic leukemia not having achieved remission |
| C94.31 | ICD10CM | Mast cell leukemia, in remission |
| C91.42 | ICD10CM | Hairy cell leukemia, in relapse |
| C94.80 | ICD10CM | Other specified leukemias not having achieved remission |
| C94.22 | ICD10CM | Acute megakaryoblastic leukemia, in relapse |
| C95.90 | ICD10CM | Leukemia, unspecified not having achieved remission |
| C95.10 | ICD10CM | Chronic leukemia of unspecified cell type not having achieved remission |
| C94.01 | ICD10CM | Acute erythroid leukemia, in remission |
| C94.82 | ICD10CM | Other specified leukemias, in relapse |
| C95.91 | ICD10CM | Leukemia, unspecified, in remission |

Table S2 Continued. ICD 9/10 CM codes used for identification of patients with leukemia.

| Code | Version | Description |
|---|---|---|
| C95.02 | ICD10CM | Acute leukemia of unspecified cell type, in relapse |
| C91.41 | ICD10CM | Hairy cell leukemia, in remission |
| C94.02 | ICD10CM | Acute erythroid leukemia, in relapse |
| C94.81 | ICD10CM | Other specified leukemias, in remission |
| C91.40 | ICD10CM | Hairy cell leukemia not having achieved remission |
| C94.00 | ICD10CM | Acute erythroid leukemia, not having achieved remission |
| C95.9 | ICD10CM | Leukemia, unspecified |
| C95.1 | ICD10CM | Chronic leukemia of unspecified cell type |
| C94.21 | ICD10CM | Acute megakaryoblastic leukemia, in remission |
| C95.92 | ICD10CM | Leukemia, unspecified, in relapse |
| C95.12 | ICD10CM | Chronic leukemia of unspecified cell type, in relapse |
| C95.00 | ICD10CM | Acute leukemia of unspecified cell type not having achieved remission |
| C94.3 | ICD10CM | Mast cell leukemia |
| C90.10 | ICD10CM | Plasma cell leukemia not having achieved remission |
| C95.11 | ICD10CM | Chronic leukemia of unspecified cell type, in remission |
| C94.2 | ICD10CM | Acute megakaryoblastic leukemia |
| C91.4 | ICD10CM | Hairy cell leukemia |
| C95.01 | ICD10CM | Acute leukemia of unspecified cell type, in remission |
| C94.32 | ICD10CM | Mast cell leukemia, in relapse |
| C94.0 | ICD10CM | Acute erythroid leukemia |
| C90.11 | ICD10CM | Plasma cell leukemia in remission |
| C90.1 | ICD10CM | Plasma cell leukemia |
| C94.8 | ICD10CM | Other specified leukemias |
| C90.12 | ICD10CM | Plasma cell leukemia in relapse |
| C91.1 | ICD10CM | Chronic lymphocytic leukemia of B-cell type |
| C91.10 | ICD10CM | Chronic lymphocytic leukemia of B-cell type not having achieved remission |
| C91.12 | ICD10CM | Chronic lymphocytic leukemia of B-cell type in relapse |
| C91.11 | ICD10CM | Chronic lymphocytic leukemia of B-cell type in remission |
| C93.00 | ICD10CM | Acute monoblastic/monocytic leukemia, not having achieved remission |
| C93.0 | ICD10CM | Acute monoblastic/monocytic leukemia |
| C93.90 | ICD10CM | Monocytic leukemia, unspecified, not having achieved remission |
| C93.9 | ICD10CM | Monocytic leukemia, unspecified |
| C93.Z2 | ICD10CM | Other monocytic leukemia, in relapse |
| C93.3 | ICD10CM | Juvenile myelomonocytic leukemia |
| C93.92 | ICD10CM | Monocytic leukemia, unspecified in relapse |
| C93.12 | ICD10CM | Chronic myelomonocytic leukemia, in relapse |
| C93.10 | ICD10CM | Chronic myelomonocytic leukemia not having achieved remission |
| C93.91 | ICD10CM | Monocytic leukemia, unspecified in remission |
| C93.Z | ICD10CM | Other monocytic leukemia |
| C93.11 | ICD10CM | Chronic myelomonocytic leukemia, in remission |
| C93.32 | ICD10CM | Juvenile myelomonocytic leukemia, in relapse |
| C93.31 | ICD10CM | Juvenile myelomonocytic leukemia, in remission |
| C93.30 | ICD10CM | Juvenile myelomonocytic leukemia, not having achieved remission |
| C93.01 | ICD10CM | Acute monoblastic/monocytic leukemia, in remission |

Table S2 Continued. ICD 9/10 CM codes used for identification of patients with leukemia.

| Code | Version | Description |
|---|---|---|
| C93.Z1 | ICD10CM | Other monocytic leukemia, in remission |
| C93 | ICD10CM | Monocytic leukemia |
| C93.Z0 | ICD10CM | Other monocytic leukemia, not having achieved remission |
| C93.02 | ICD10CM | Acute monoblastic/monocytic leukemia, in relapse |
| C93.1 | ICD10CM | Chronic myelomonocytic leukemia |
| C92.01 | ICD10CM | Acute myeloblastic leukemia, in remission |
| C92.A1 | ICD10CM | Acute myeloid leukemia with multilineage dysplasia, in remission |
| C92.4 | ICD10CM | Acute promyelocytic leukemia |
| C92.41 | ICD10CM | Acute promyelocytic leukemia, in remission |
| C92.60 | ICD10CM | Acute myeloid leukemia with 11q23-abnormality not having achieved remission |
| C92.42 | ICD10CM | Acute promyelocytic leukemia, in relapse |
| C92.A | ICD10CM | Acute myeloid leukemia with multilineage dysplasia |
| C92.5 | ICD10CM | Acute myelomonocytic leukemia |
| C92.A0 | ICD10CM | Acute myeloid leukemia with multilineage dysplasia, not having achieved remission |
| C92.62 | ICD10CM | Acute myeloid leukemia with 11q23-abnormality in relapse |
| C92.51 | ICD10CM | Acute myelomonocytic leukemia, in remission |
| C92.0 | ICD10CM | Acute myeloblastic leukemia |
| C92.6 | ICD10CM | Acute myeloid leukemia with 11q23-abnormality |
| C92.40 | ICD10CM | Acute promyelocytic leukemia, not having achieved remission |
| C92.50 | ICD10CM | Acute myelomonocytic leukemia, not having achieved remission |
| C92.52 | ICD10CM | Acute myelomonocytic leukemia, in relapse |
| C92.A2 | ICD10CM | Acute myeloid leukemia with multilineage dysplasia, in relapse |
| C92.61 | ICD10CM | Acute myeloid leukemia with 11q23-abnormality in remission |
| C92.02 | ICD10CM | Acute myeloblastic leukemia, in relapse |
| C92.00 | ICD10CM | Acute myeloblastic leukemia, not having achieved remission |
| C90.20 | ICD10CM | Extramedullary plasmacytoma not having achieved remission |
| C90.30 | ICD10CM | Solitary plasmacytoma not having achieved remission |
| C90.2 | ICD10CM | Extramedullary plasmacytoma |
| C90.00 | ICD10CM | Multiple myeloma not having achieved remission |
| C88.9 | ICD10CM | Malignant immunoproliferative disease, unspecified |
| C90.01 | ICD10CM | Multiple myeloma in remission |
| C90.0 | ICD10CM | Multiple myeloma |
| C88.3 | ICD10CM | Immunoproliferative small intestinal disease |
| C90.31 | ICD10CM | Solitary plasmacytoma in remission |
| C90.02 | ICD10CM | Multiple myeloma in relapse |
| C90.21 | ICD10CM | Extramedullary plasmacytoma in remission |
| C88.2 | ICD10CM | Heavy chain disease |
| C90.22 | ICD10CM | Extramedullary plasmacytoma in relapse |
| C90.32 | ICD10CM | Solitary plasmacytoma in relapse |
| C91.Z | ICD10CM | Other lymphoid leukemia |
| C91.62 | ICD10CM | Prolymphocytic leukemia of T-cell type, in relapse |
| C91.52 | ICD10CM | Adult T-cell lymphoma/leukemia (HTLV-1-associated), in relapse |
| C91.92 | ICD10CM | Lymphoid leukemia, unspecified, in relapse |
| C91.A2 | ICD10CM | Mature B-cell leukemia Burkitt-type, in relapse |

Table S2 Continued. ICD 9/10 CM codes used for identification of patients with leukemia.

| Code | Version | Description |
|---|---|---|
| C91.6 | ICD10CM | Prolymphocytic leukemia of T-cell type |
| C91.31 | ICD10CM | Prolymphocytic leukemia of B-cell type, in remission |
| C91.61 | ICD10CM | Prolymphocytic leukemia of T-cell type, in remission |
| C91.90 | ICD10CM | Lymphoid leukemia, unspecified not having achieved remission |
| C91.A0 | ICD10CM | Mature B-cell leukemia Burkitt-type not having achieved remission |
| C91.9 | ICD10CM | Lymphoid leukemia, unspecified |
| C91.5 | ICD10CM | Adult T-cell lymphoma/leukemia (HTLV-1-associated) |
| C91.91 | ICD10CM | Lymphoid leukemia, unspecified, in remission |
| C91.30 | ICD10CM | Prolymphocytic leukemia of B-cell type not having achieved remission |
| C91.Z2 | ICD10CM | Other lymphoid leukemia, in relapse |
| C91 | ICD10CM | Lymphoid leukemia |
| C91.Z0 | ICD10CM | Other lymphoid leukemia not having achieved remission |
| C91.3 | ICD10CM | Prolymphocytic leukemia of B-cell type |
| C91.Z1 | ICD10CM | Other lymphoid leukemia, in remission |
| C91.51 | ICD10CM | Adult T-cell lymphoma/leukemia (HTLV-1-associated), in remission |
| C91.50 | ICD10CM | Adult T-cell lymphoma/leukemia (HTLV-1-associated) not having achieved remission |
| C91.60 | ICD10CM | Prolymphocytic leukemia of T-cell type not having achieved remission |
| C91.A1 | ICD10CM | Mature B-cell leukemia Burkitt-type, in remission |
| C91.32 | ICD10CM | Prolymphocytic leukemia of B-cell type, in relapse |

Table S3. CPT, ICD-10-PCS, and ICD-9-Proc codes used to calculate assisted ventilation days.

| Code | Version | Description |
|---|---|---|
| 94656 | CPT4 | Ventilation assist and management, initiation of pressure or volume preset ventilators for assisted or controlled breathing; first day |
| 94657 | CPT4 | Ventilation assist and management, initiation of pressure or volume preset ventilators for assisted or controlled breathing; subsequent days |
| 94004 | CPT4 | Ventilation assist and management, initiation of pressure or volume preset ventilators for assisted or controlled breathing; nursing facility, per day |
| 94003 | CPT4 | Ventilation assist and management, initiation of pressure or volume preset ventilators for assisted or controlled breathing; hospital inpatient/observation, each subsequent day |
| 94002 | CPT4 | Ventilation assist and management, initiation of pressure or volume preset ventilators for assisted or controlled breathing; hospital inpatient/observation, initial day |
| 5A1945Z | ICD-10-PCS | Respiratory Ventilation, 24-96 Consecutive Hours |
| 5A1955Z | ICD-10-PCS | Respiratory Ventilation, Greater than 96 Consecutive Hours |
| 5A1935Z | ICD-10-PCS | Respiratory Ventilation, Less than 24 Consecutive Hours |
| 96.7 | ICD-9-Proc | Continuous invasive mechanical ventilation of unspecified duration |
| 96.71 | ICD-9-Proc | Continuous invasive mechanical ventilation for less than 96 consecutive hours |
| 96.72 | ICD-9-Proc | Continuous invasive mechanical ventilation for 96 consecutive hours or more |

Table S4. CPT, ICD-10-PCS, and ICD-9-Proc codes used to calculate dialysis days.

| code | version | Description |
|---|---|---|
| 39.95 | ICD9Proc | Hemodialysis |
| 38.95 | ICD9Proc | Venous catheterization for renal dialysis |
| 5A1D00Z | ICD10PCS | Performance of Urinary Filtration, Single (Deprecated) |
| 5A1D90Z | ICD10PCS | Performance of Urinary Filtration, Continuous, Greater than 18 hours Per Day |
| 5A1D70Z | ICD10PCS | Performance of Urinary Filtration, Intermittent, Less than 6 Hours Per Day |
| 5A1D80Z | ICD10PCS | Performance of Urinary Filtration, Prolonged Intermittent, 6-18 hours Per Day |
| 90947 | CPT4 | Dialysis procedure other than hemodialysis, requiring repeated physician evaluations. |
| 90945 | CPT4 | Dialysis procedure other than hemodialysis (e.g., peritoneal dialysis, hemofiltration, or other continuous renal replacement therapies), with a single physician evaluation. |
| 90935 | CPT4 | Hemodialysis procedure with a single evaluation by a physician or other qualified healthcare professional. |
| 90937 | CPT4 | Hemodialysis procedure requiring repeated evaluation(s) with or without substantial revision of dialysis prescription. |
| 36148 | CPT4 | Introduction of needle or intracatheter; arteriovenous shunt created for dialysis (cannula, f istula, or graft) (Deprecated) |
| 36147 | CPT4 | Introduction of needle and/or catheter, arteriovenous shunt created for dialysis (graft/fistula); initial access with complete radiological evaluation of dialysis access, including fluoroscopy, image documentation, and report. |
| 36800 | CPT4 | Insertion of cannula for hemodialysis, other purpose (separate procedure); vein to vein |
| 90997 | CPT4 | Hemoperfusion |

Table S5. ICD 9/10 codes used to identify patients with end stage renal disease

| Acute Renal Failure Codes | | |
|---|---|---|
| Code | Version | Description |
| 585.5 | ICD9CM | Chronic kidney disease, Stage V |
| 585.6 | ICD9CM | End stage renal disease |
| N18.6 | ICD10CM | End stage renal disease |
| N18.5 | ICD10CM | Chronic kidney disease, stage 5 |

Table S6. ICU sepsis patient cohort Elixhauser comorbidities.

| | No CHIP N = 3,010[1] | CHIP N = 220[1] | Age-Adjusted P[2] |
|---|---|---|---|
| **Comorbidities** | | | |
| Congestive Heart Failure | 842 (28%) | 72 (33%) | 0.69 |
| Arrhythmia | 2,302 (77%) | 182 (83%) | 0.25 |
| Valvular Disease | 631 (21%) | 48 (22%) | 0.42 |
| Pulm. Circulatory Disorders | 505 (17%) | 31 (14%) | 0.39 |
| Peripheral Vascular Disease | 486 (16%) | 59 (27%) | 0.1 |
| Uncomplicated Hypertension | 1,571 (52%) | 122 (55%) | 0.13 |
| Complicated Hypertension | 458 (15%) | 37 (17%) | 0.66 |
| Paralysis | 174 (5.8%) | 10 (4.5%) | 0.83 |
| Other Neurological Disorders | 688 (23%) | 56 (25%) | 0.47 |
| Chronic Pulm. Disease | 947 (31%) | 82 (37%) | 0.71 |
| Uncomplicated Diabetes | 720 (24%) | 73 (33%) | 0.09 |
| Complicated Diabetes | 437 (15%) | 25 (11%) | 0.05 |
| Hypothyroidism | 472 (16%) | 39 (18%) | 0.5 |
| Renal Failure | 812 (27%) | 65 (30%) | 0.82 |
| Liver Disease | 753 (25%) | 64 (29%) | 0.06 |
| Peptic Ulcer Disease | 63 (2.1%) | 9 (4.1%) | 0.05 |
| AIDS/HIV | 52 (1.7%) | 2 (0.9%) | 0.82 |
| Lymphoma | 78 (2.6%) | 4 (1.8%) | 0.35 |
| Metastatic Cancer | 309 (10%) | 15 (6.8%) | 0.07 |
| Solid Tumor | 934 (31%) | 85 (39%) | 0.37 |
| Rheumatoid Arthritis | 197 (6.6%) | 15 (6.8%) | 0.95 |
| Coagulopathy | 1,077 (36%) | 83 (38%) | 0.61 |
| Obesity | 513 (17%) | 24 (11%) | 0.06 |
| Weight Loss | 1,034 (34%) | 84 (38%) | 0.48 |
| Fluid/Electrolyte Disorder | 2,417 (80%) | 179 (81%) | 0.88 |
| Blood Loss Anemia | 135 (4.5%) | 13 (5.9%) | 0.13 |
| Deficiency Anemia | 795 (26%) | 60 (27%) | 0.42 |
| Alcohol Abuse | 639 (21%) | 44 (20%) | 0.84 |
| Drug Abuse | 152 (5.1%) | 7 (3.2%) | 0.97 |
| Psychoses | 121 (4.0%) | 10 (4.5%) | 0.29 |
| Depression | 625 (21%) | 44 (20%) | 0.49 |
| Elixhauser Comorbidity Score | 20 (13, 28) | 22 (15, 30) | 0.26 |
| Elixhauser Number of Comorbidities | 7 (5, 9) | 7 (6, 9) | 0.43 |

[1] Median (Q1, Q3); n (%)
[2] Binary Logistic or Linear regression

Table S7. ICU non-sepsis cohort patient characteristics.

| | No CHIP N = 1,991[1] | CHIP N = 130[1] | p-value[2] | q-value[3] |
|---|---|---|---|---|
| **Gender** | | | >0.9 | >0.9 |
| Female | 881 (44%) | 57 (44%) | | |
| **Age at Admission** | 58 (47, 68) | 67 (59, 75) | <0.001 | <0.001 |
| **Admitting ICU** | | | 0.4 | 0.5 |
| Cardiovascular | 652 (33%) | 44 (34%) | | |
| Neurological | 613 (31%) | 34 (26%) | | |
| Trauma | 86 (4.3%) | 3 (2.3%) | | |
| Surgical | 640 (32%) | 49 (38%) | | |
| **Race** | | | 0.2 | 0.4 |
| Black | 243 (12%) | 11 (8.5%) | | |
| Other | 47 (2.4%) | 1 (0.8%) | | |
| White | 1,701 (85%) | 118 (91%) | | |
| **Body Mass Index** | 28 (24, 33) | 27 (23, 31) | 0.003 | 0.008 |
| **Baseline Measures** | | | | |
| Maximum Bilirubin, mg/dL | 0.9 (0.5, 1.9) | 0.8 (0.6, 1.4) | 0.6 | 0.8 |
| Maximum Creatinine, mg/dL | 1.0 (0.8, 1.3) | 1.0 (0.8, 1.4) | 0.3 | 0.7 |
| Minimum Platelets, $10^3$/μL | 180 (124, 236) | 167 (118, 226) | 0.4 | 0.7 |
| Minimum Systolic Blood Pressure, mmHg | 96 (87, 107) | 97 (86, 107) | 0.9 | >0.9 |
| Maximum Respiratory Rate, per minute | 24 (21, 27) | 24 (21, 26) | 0.6 | 0.8 |
| Minimum $SpO_2$,% | 93 (91, 95) | 93 (90, 95) | 0.2 | 0.7 |
| Minimum $PaO_2$:$FiO_2$ Ratio | 251(165, 340) | 253 (195, 355) | 0.3 | 0.7 |
| Maximum Pulse Rate, per minute | 101 (90, 115) | 101 (90, 113) | 0.9 | >0.9 |
| **Vasopressors at Baseline** | 476 (23.9%) | 26 (20.0%) | 0.3 | 0.7 |
| **Mechanical Ventilation at Baseline** | 610 (30.6%) | 33 (25.4%) | 0.2 | 0.7 |
| **SOFA Organ System Scores** | | | | |
| Respiratory | 1.0 (1.0, 3.0) | 1.0 (1.0, 3.0) | 0.4 | 0.7 |
| Renal | 0.0 (0.0, 1.0) | 0.0 (0.0, 1.0) | 0.14 | 0.7 |
| Hepatic | 0.0 (0.0, 1.0) | 0.0 (0.0, 1.0) | 0.6 | 0.8 |
| Coagulation | 0.0 (0.0, 1.0) | 0.0 (0.0, 1.0) | 0.5 | 0.8 |
| Cardiovascular | 1.0 (0.0, 1.0) | 1.0 (0.0, 1.0) | 0.8 | >0.9 |
| **Total SOFA Score** | 3.0 (2.0, 6.0) | 3.0 (2.0, 5.0) | 0.8 | >0.9 |

[1] N (%); Median (Q1, Q3)

[2] Pearson's Chi-squared test; Wilcoxon rank sum test

[3] False discovery rate correction for multiple testing

Table S8. ICU non-sepsis patient cohort Elixhauser comorbidities.

| | No CHIP N = 1,991[1] | CHIP N = 130[1] | Age-Adjusted P[2] |
|---|---|---|---|
| **Comorbidities** | | | |
| Congestive Heart Failure | 479 (24%) | 32 (25%) | 0.370 |
| Arrhythmia | 945 (48%) | 73 (57%) | 0.717 |
| Valvular Disease | 444 (23%) | 33 (26%) | 0.819 |
| Pulm. Circulatory Disorders | 228 (12%) | 14 (11%) | 0.685 |
| Peripheral Vascular Disease | 296 (15%) | 27 (21%) | 0.837 |
| Uncomplicated Hypertension | 1,175 (60%) | 95 (74%) | 0.163 |
| Complicated Hypertension | 199 (10%) | 18 (14%) | 0.715 |
| Paralysis | 93 (4.7%) | 9 (7.0%) | 0.378 |
| Other Neurological Disorders | 297 (15%) | 22 (17%) | 0.426 |
| Chronic Pulm. Disease | 495 (25%) | 41 (32%) | 0.434 |
| Uncomplicated Diabetes | 441 (23%) | 29 (22%) | 0.285 |
| Complicated Diabetes | 189 (9.6%) | 12 (9.3%) | 0.501 |
| Hypothyroidism | 274 (14%) | 17 (13%) | 0.396 |
| Renal Failure | 313 (16%) | 24 (19%) | 0.969 |
| Liver Disease | 217 (11%) | 11 (8.5%) | 0.324 |
| Peptic Ulcer Disease | 23 (1.2%) | 1 (0.8%) | 0.622 |
| AIDS/HIV | 10 (0.5%) | 2 (1.6%) | 0.050 |
| Lymphoma | 12 (0.6%) | 0 (0%) | 0.988 |
| Metastatic Cancer | 137 (7.0%) | 7 (5.4%) | 0.330 |
| Solid Tumor | 523 (27%) | 34 (26%) | 0.291 |
| Rheumatoid Arthritis | 63 (3.2%) | 8 (6.2%) | 0.091 |
| Coagulopathy | 367 (19%) | 30 (23%) | 0.420 |
| Obesity | 361 (18%) | 12 (9.3%) | 0.028 |
| Weight Loss | 305 (16%) | 17 (13%) | 0.209 |
| Fluid/Electrolyte Disorder | 1,048 (53%) | 66 (51%) | 0.170 |
| Blood Loss Anemia | 39 (2.0%) | 4 (3.1%) | 0.251 |
| Deficiency Anemia | 271 (14%) | 20 (16%) | 0.859 |
| Alcohol Abuse | 377 (19%) | 25 (19%) | 0.655 |
| Drug Abuse | 49 (2.5%) | 5 (3.9%) | 0.171 |
| Psychoses | 55 (2.8%) | 2 (1.6%) | 0.615 |
| Depression | 339 (17%) | 16 (12%) | 0.277 |
| Elixhauser Comorbidity Score | 12 (5, 20) | 12 (7, 21) | 0.390 |
| Elixhauser Number of Comorbidities | 5.00 (3.00, 7.00) | 5.00 (4.00, 7.00) | 0.507 |

[1] Median (Q1, Q3); n (%)
[2] Binary Logistic or Linear regression

Table S9. Logistic regression model results for odds of inpatient mortality in critically-ill sepsis patients.

| | OR | 95% CI | P-value[1] |
|---|---|---|---|
| **CHIP Status** | | | |
| No CHIP | — | — | |
| CHIP | 1.54 | 1.13, 2.07 | 0.005 |
| **Gender** | | | |
| Female | — | — | |
| Male | 0.97 | 0.81, 1.16 | 0.7 |
| **Total SOFA Score** | 1.21 | 1.18, 1.24 | <0.001 |
| **Age at Admission** | 1.01 | 1.01, 1.02 | <0.001 |
| **Race** | | | |
| Black | — | — | |
| Other | 1.44 | 0.86, 2.36 | 0.2 |
| White | 1.08 | 0.84, 1.41 | 0.5 |
| **CHIP Status * Age at Admission** | 1.00 | 0.98, 1.03 | >0.9 |
| **CHIP Status * Total SOFA Score** | 1.01 | 0.91, 1.13 | 0.9 |

OR = Odds Ratio, CI = Confidence Interval
[1]Wald's t-test

Table S10. Survival probability in sepsis survivors with and without clonal hematopoiesis of indeterminant potential (CHIP).

| | Survival Probability (95% CI) | | |
|---|---|---|---|
| **Survival at** | **CHIP** | **No CHIP** | ***P value*[1]** |
| 1 year | 0.55 (0.45, 0.70) | 0.75 (0.72, 0.77) | < 0.001 |
| 2 years | 0.46 (0.35, 0.60) | 0.69 (0.66, 0.72) | 0.049 |
| 5 years | 0.39 (0.28, 0.54) | 0.60 (0.57, 0.63) | 0.037 |

[1]Z-test

Table S11. Case-Control matching results for variant allele fraction (VAF) growth analysis

| | **Control**<br>N = 22[1] | **Sepsis**<br>N = 11[1] | **p-value**[2] |
|---|---|---|---|
| **Sex** | | | 0.8 |
| Female | 11 (50%) | 6 (55%) | |
| Male | 11 (50%) | 5 (45%) | |
| **Age at First Sample** | 70.0 (68.0, 78.0) | 73.0 (70.0, 75.0) | 0.5 |
| **Allele Frequency at first Sample** | 0.06 (0.03, 0.11) | 0.07 (0.02, 0.13) | >0.9 |
| **Gene** | | | >0.9 |
| DNMT3A | 6 (27%) | 3 (27%) | |
| PPM1D | 6 (27%) | 3 (27%) | |
| SF3B1 | 2 (9.1%) | 1 (9.1%) | |
| TET2 | 4 (18%) | 2 (18%) | |
| TP53 | 4 (18%) | 2 (18%) | |
| **Mutation Type** | | | >0.9 |
| frameshift deletion | 2 (9.1%) | 1 (9.1%) | |
| nonsynonymous SNV | 14 (64%) | 7 (64%) | |
| Stop-gain | 6 (27%) | 3 (27%) | |
| **Time interval Between first and second sample (y)** | 4.45 (4.00, 5.30) | 4.60 (4.00, 4.60) | 0.7 |
| **Race** | | | 0.5 |
| Black | 3 (14%) | 0 (0%) | |
| White | 19 (86%) | 11 (100%) | |

[1] n (%); Median (Q1, Q3)

Table S12. Blood counts and differentials matched to first and second specimen collections used to measure CHIP variant allele fraction growth.

| | First Bio-Specimen | | | Second Bio-Specimen | | |
|---|---|---|---|---|---|---|
| **Measure**[1] | **Control**[2] | **Sepsis**[2] | **p-value**[3] | **Control**[2] | **Sepsis**[2] | **p-value**[3] |
| White Blood Count | 6.40 (3.20, 8.25) | 7.10 (5.04, 8.40) | 0.8 | 5.0 (5.0, 8.0) | 12.0 (10.0, 17.0) | 0.065 |
| Neutrophil Count | 4.18 (1.68, 5.65) | 4.89 (4.61, 5.04) | >0.9 | 4.0 (3.0, 6.0) | 10.0 (6.0, 15.0) | 0.093 |
| Relative Neutrophils | 0.61 (0.50, 0.64) | 0.69 (0.60, 0.91) | 0.3 | 0.67 (0.56, 0.75) | 0.82 (0.66, 0.90) | 0.13 |
| Lymphocyte Count | 1.55 (0.99, 2.06) | 1.58 (0.37, 1.59) | 0.5 | 1.28 (1.25, 1.52) | 0.74 (0.58, 1.66) | 0.3 |
| Relative Lymphocyte | 0.27 (0.23, 0.36) | 0.19 (0.07, 0.22) | 0.085 | 0.21 (0.15, 0.34) | 0.10 (0.04, 0.22) | 0.093 |
| Monocyte Count | 1.55 (0.99, 2.06) | 1.58 (0.37, 1.59) | 0.5 | 1.28 (1.25, 1.52) | 0.74 (0.58, 1.66) | 0.3 |
| Relative Monocyte | 0.09 (0.08, 0.10) | 0.05 (0.01, 0.17) | 0.6 | 0.077 (0.073, 0.081) | 0.061 (0.037, 0.094) | 0.3 |

[1] Cell counts in $10^3$/microliter. Percentages are represented as fractions.

[2] Median (Q1, Q3)

[3] Wilcoxon rank sum test

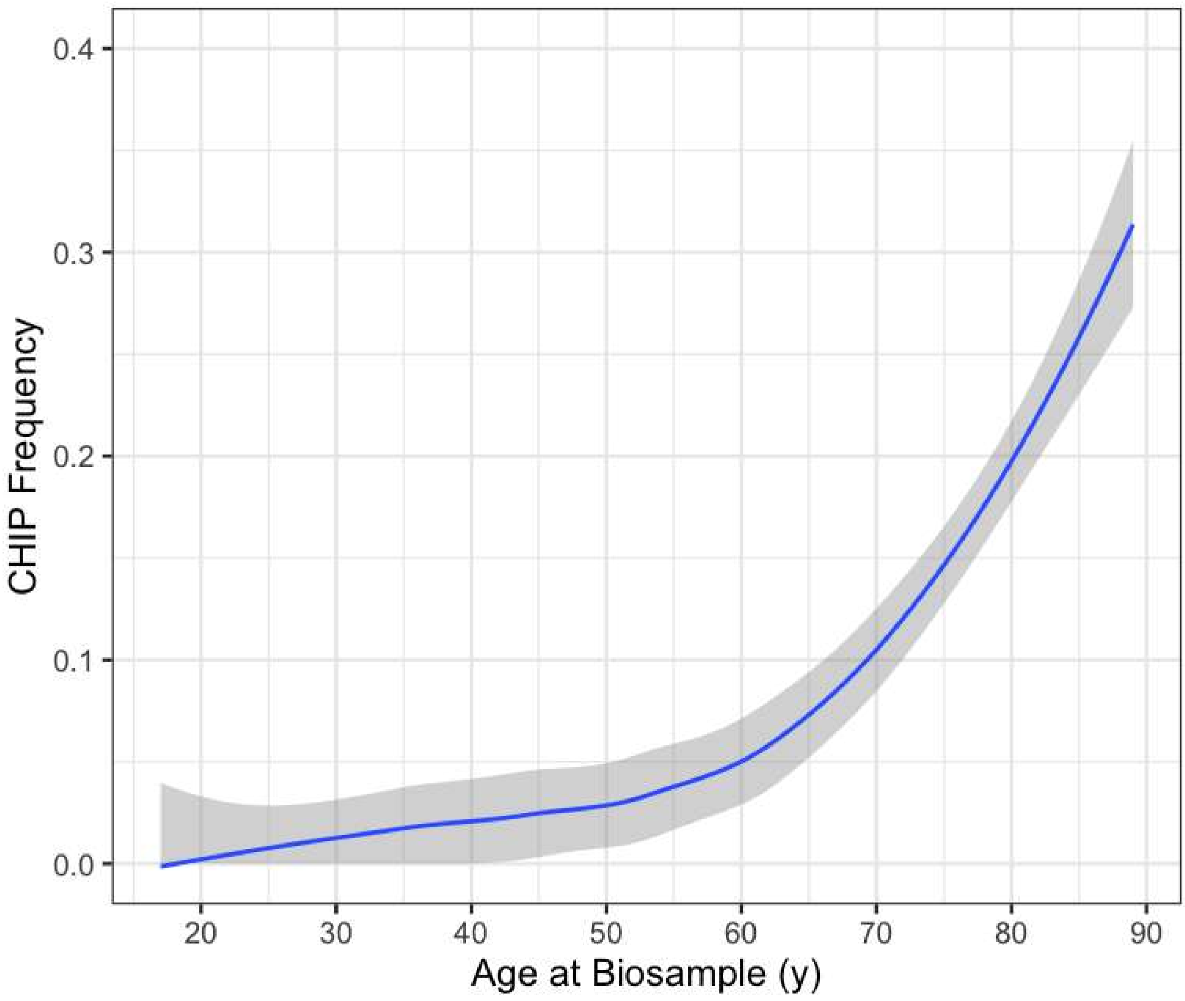


Figure S1. Distribution of CHIP fraction across age in patients admitted to the ICU. The blue line represents the fitted Locally Estimated Scatterplot Smoothing (LOESS) regression. Gray shading represents 95% confidence intervals.

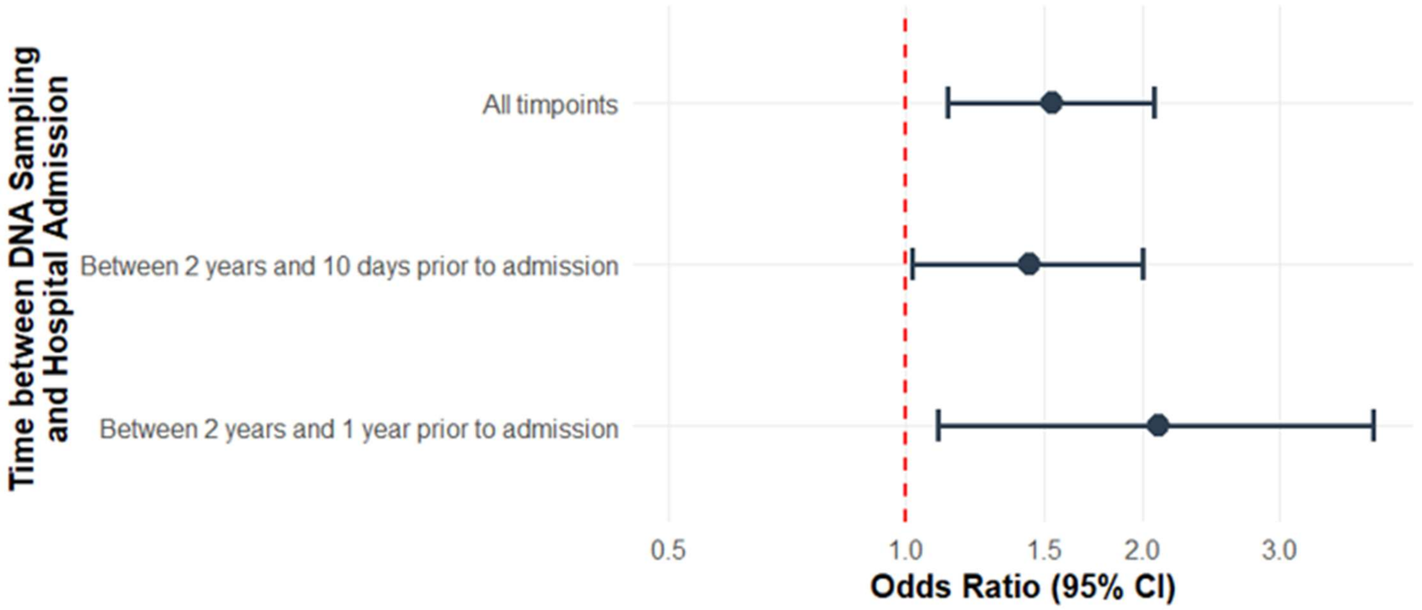


Figure S2. Sensitivity analysis of ICU sepsis mortality by DNA collection timing. Intervals were based on filtering out DNA samples that were collected from patients within 10 days or 1 year prior to their hospital admission. Mortality is predicted by logistic regression, with Odds Ratio representing risk for mortality in patients with clonal hematopoiesis of indeterminant potential (CHIP) versus those without. Data are adjusted for sex, race, age at time of admission, and baseline sequential organ failure assessment (SOFA) scores. Bars represent 95% confidence intervals.

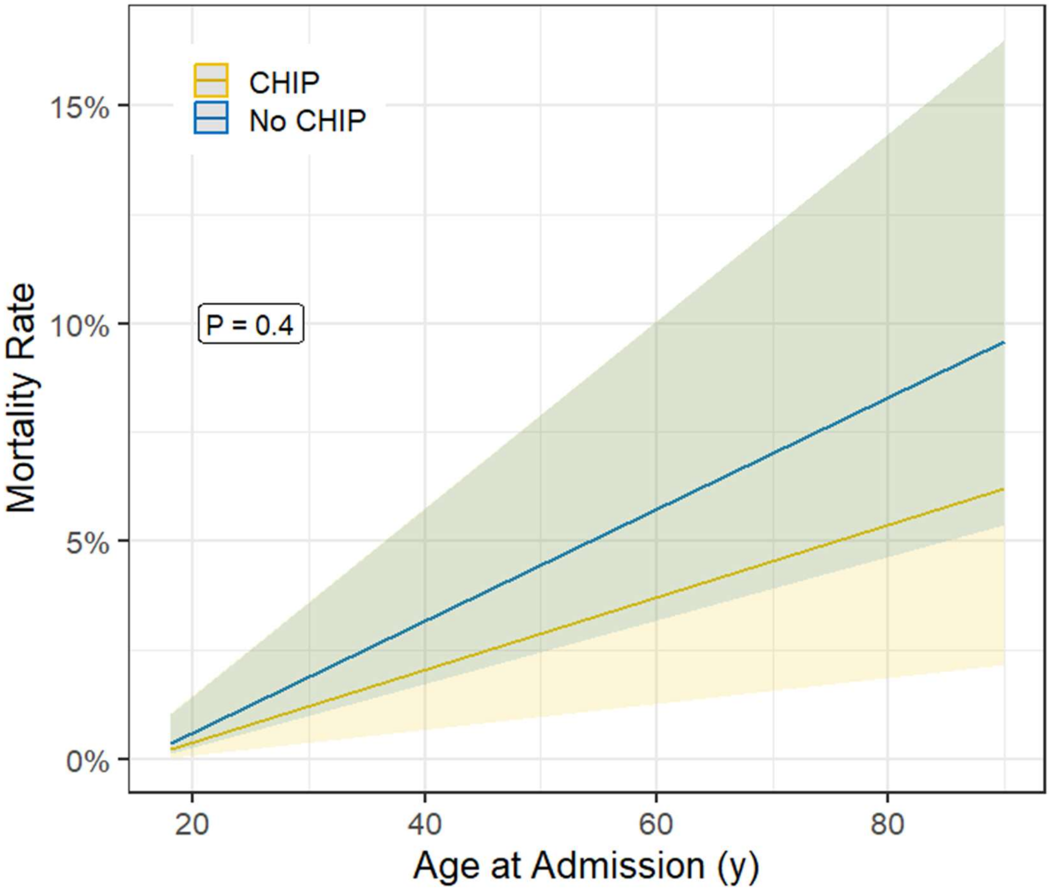


Figure S3. Inpatient mortality in non-sepsis patients admitted to non-medical intensive care units. Predicted mortality by logistic regression for patients with and without clonal hematopoiesis of indeterminant potential (CHIP) after adjusting for sex, race, age at time of admission, and baseline sequential organ failure assessment (SOFA) scores. Shading represents 95% confidence intervals.

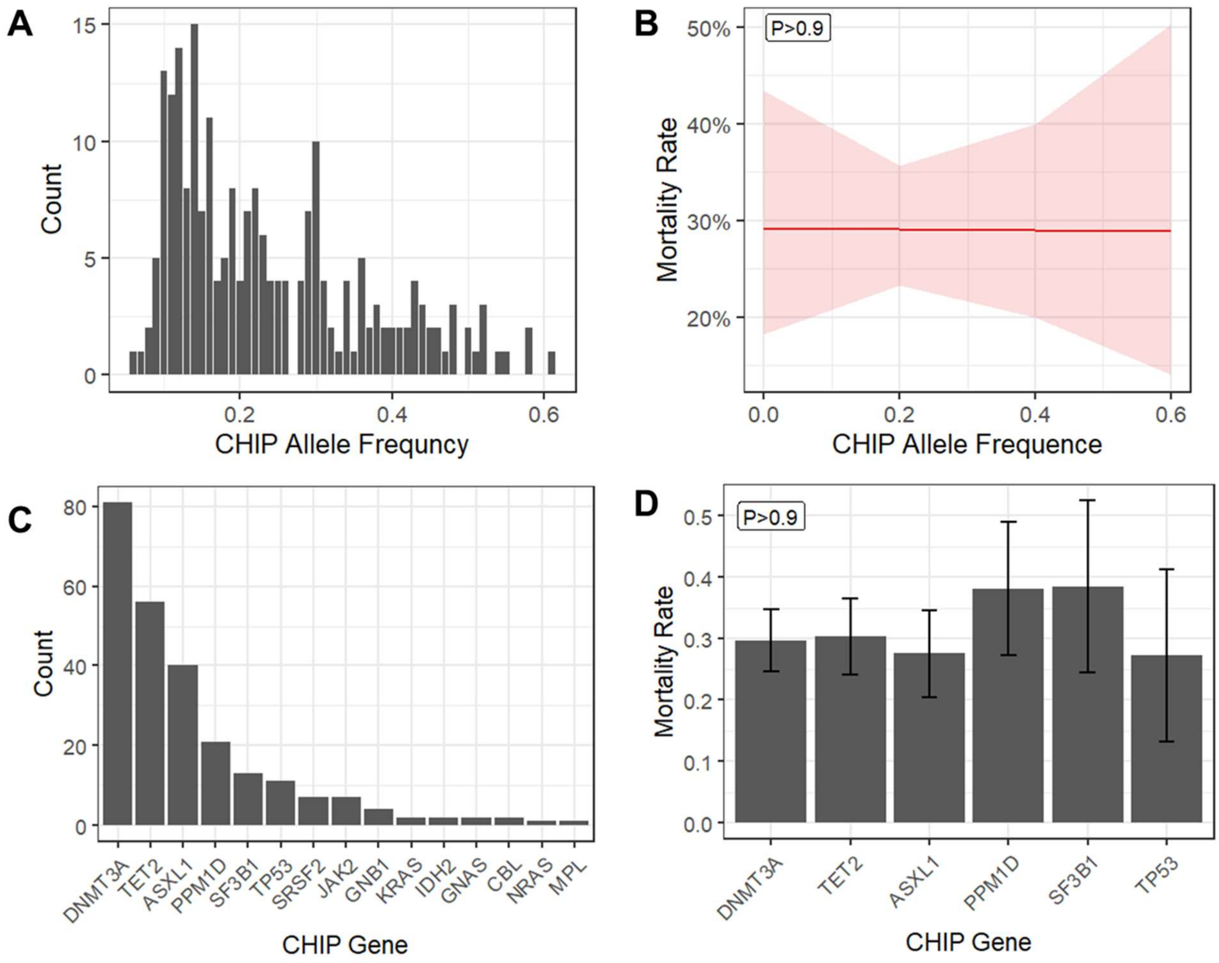


Figure S4. Association of CHIP variant allele frequency and specific CHIP gene mutation with hospital mortality in sepsis patients admitted to the ICU. A) Histogram of CHIP allele fraction in all patients with detectable CHIP. B) Predicted mortality versus CHIP allele fraction corrected for sex, race, age at time of admission and initial SOFA score (Wald Test). C) Histogram of mutated CHIP genes in patients with detectable CHIP. D) Predicted mortality versus the detectable mutated CHIP gene (one-way ANOVA).

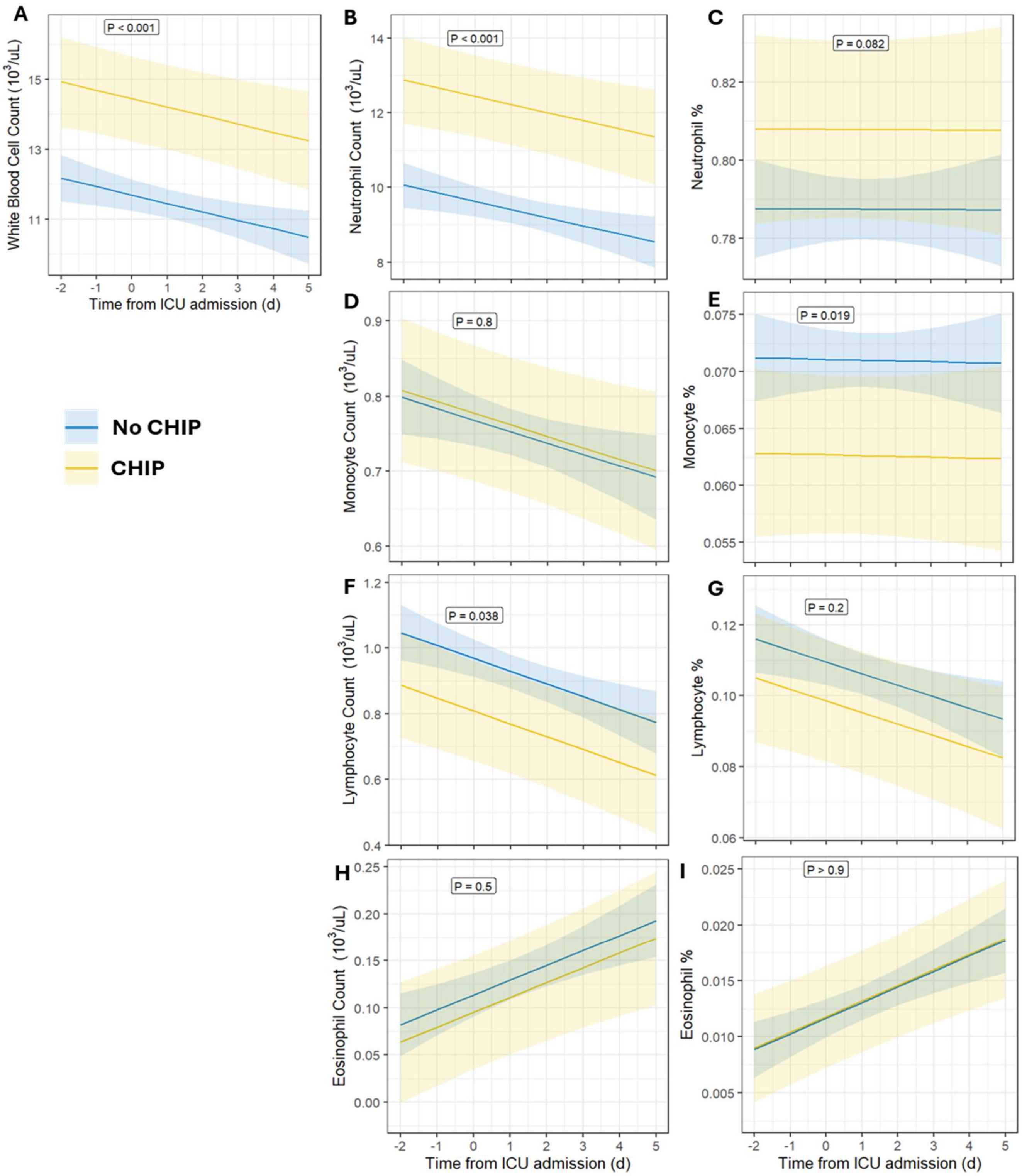


Figure S5. Association of clonal hematopoiesis of indeterminant potential (CHIP) with cell-count differentials. Regression lines from peripheral leukocyte count and differentials collected during routine care between 2 days prior to ICU admission and 5 days after admission are plotted for A) total white blood cell count, B, C) neutrophil count and percentage, D, E) monocyte count and percentage, F, G) lymphocyte count and percentage, and H, I) eosinophil count and percentage. Shading represents 95% confidence interval. P values represent results of linear regression Wald t-test after adjusting for time of blood collection relative to time of ICU admission, age at time of admission, race, initial SOFA score, and sex.

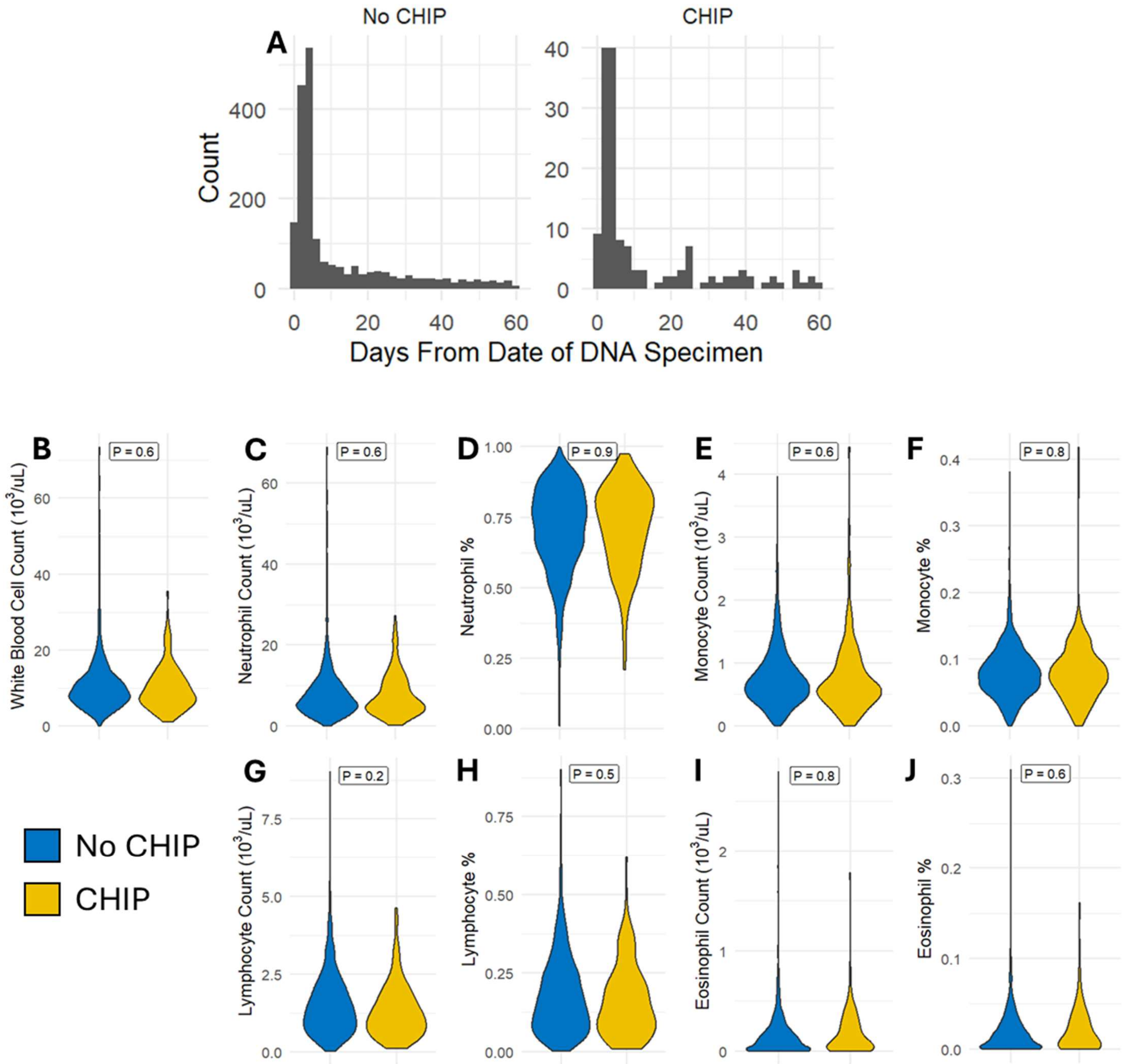


Figure S6. Leukocyte counts and differentials proximal to the time of pre-illness biospecimen collection. Leukocyte counts were used if they met two criteria: they were within 60 days of DNA biospecimen collection and occurred prior to ICU admission. A) Histograms showing the distribution of time intervals between DNA biospecimen collection and the closest complete blood count (CBC) with differential. Violin plots display absolute or percentage leukocytes for B) total white blood cells, C,D) neutrophils, E,F) monocytes, G,H) lymphocytes, and I,J) eosinophils. P values represent results of linear regression Wald t-test after adjusting for age at time of admission, race, and sex.

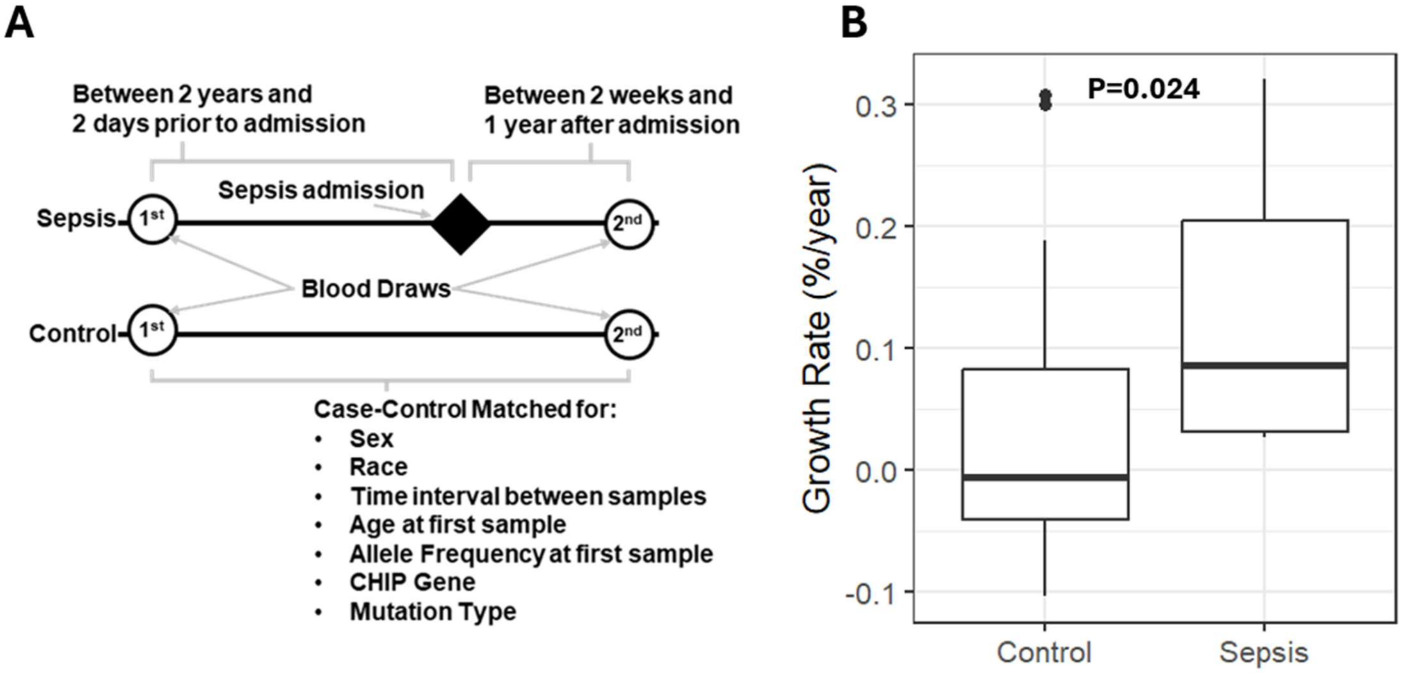


Figure S7. Clonal hematopoiesis of indeterminant potential (CHIP) variant allele fraction (VAF) expansion accelerates in sepsis patients who survive an ICU admission. A) Diagram of patient selection. Patients were selected who had two DNA samples collected; one that was collected between two years and two days prior to their admission and a second sample collected between two weeks and one year after their admission. Controls were selected in a case-control fashion, first enforcing for exact CHIP gene and mutation type, then matching for sex, race, time interval between the two samples, patient age at time of first sample draw, and allele fraction of the first sample. B) Boxplots of the growth rates for control and sepsis patients. Growth rates are calculated based on a compound interest function between two samples. P value based on linear regression Wald's t-test.

1. Arends CM, Galan-Sousa J, Hoyer K, et al (2018) Hematopoietic lineage distribution and evolutionary dynamics of clonal hematopoiesis. Leukemia 32:1908–1919. https://doi.org/10.1038/s41375-018-0047-7

2. Pershad Y, Uddin MM, Xue L, et al (2025) Correlates and consequences of clonal hematopoiesis expansion rate: a 16-year longitudinal study of 6976 women. Blood 146:1078–1087. https://doi.org/10.1182/blood.2025028417

3. Mack T, Pershad Y, Vlasschaert C, et al (2025) Germline genetics, disease, and exposure to medication influence longitudinal dynamics of clonal hematopoiesis. Haematologica 110:1010–1018. https://doi.org/10.3324/haematol.2024.286513

4. Mack T, Vlasschaert C, Beck K von, et al (2024) Cost-Effective and Scalable Clonal Hematopoiesis Assay Provides Insight into Clonal Dynamics. The Journal of Molecular Diagnostics 26:563–573. https://doi.org/10.1016/j.jmoldx.2024.03.007
